# Background incidence rates of adverse events of special interest related to COVID-19 vaccines in Ontario, Canada, 2015 to 2020, to inform COVID-19 vaccine safety surveillance

**DOI:** 10.1101/2022.01.12.22269169

**Authors:** Sharifa Nasreen, Andrew Calzavara, Sarah A. Buchan, Nisha Thampi, Caitlin Johnson, Sarah E. Wilson, Jeffrey C. Kwong, on behalf of the Canadian Immunization Research Network (CIRN) Provincial Collaborative Network (PCN) Ontario investigators

## Abstract

**Background:** Background incidence rates are critical in pharmacovigilance to facilitate identification of vaccine safety signals. We estimated background incidence rates of nine adverse events of special interest related to COVID-19 vaccines in Ontario, Canada.

**Methods:** We conducted a population-based retrospective observational study using linked health administrative databases for hospitalizations and emergency department visits among Ontario residents. We estimated incidence rates of Bell’s palsy, idiopathic thrombocytopenia, febrile convulsions, acute disseminated encephalomyelitis, myocarditis, pericarditis, Kawasaki disease, Guillain-Barré syndrome, and transverse myelitis during five pre-pandemic years (2015– 2019) and 2020.

**Results:** The average annual population was 14 million across all age groups with 51% female. The pre-pandemic mean annual rates per 100,000 population during 2015–2019 were 43.9 for idiopathic thrombocytopenia, 27.8 for Bell’s palsy, 25.0 for febrile convulsions, 22.8 for acute disseminated encephalomyelitis, 11.3 for myocarditis/pericarditis, 8.6 for pericarditis, 2.9 for myocarditis, 1.9 for Guillain-Barré syndrome, 1.7 for transverse myelitis, and 1.6 for Kawasaki disease. Females had higher rates of acute disseminated encephalomyelitis and transverse myelitis while males had higher rates of myocarditis, pericarditis, and Guillain-Barré syndrome. Bell’s palsy, acute disseminated encephalomyelitis, and Guillain-Barré syndrome increased with age. The mean rates of myocarditis and/or pericarditis increased with age up to 79 years; males had higher rates than females: from 12–59 years for myocarditis and ≥12 years for pericarditis. Febrile convulsions and Kawasaki disease were predominantly childhood diseases and generally decreased with age.

**Conclusions:** Our estimated background rates will permit estimating numbers of expected events for these conditions and facilitate detection of potential safety signals following COVID-19 vaccination.

## Introduction

Health Canada initially authorized BNT162b2 (Pfizer-BioNTech Comirnaty) and mRNA-1273 (Moderna SpikeVax) COVID-19 vaccines for use in individuals aged ≥16 and ≥18 years, respectively, in December 2020.^1^ The authorization was later expanded for use of BNT162b2 in adolescents aged 12–15 years in May 2021, and for use of mRNA-1273 in 12–17 years in August 2021.^2^ Health Canada authorized adenovirus vector-based ChAdOx1 COVID-19 vaccines (AstraZeneca Vaxzevria and COVISHIELD) in February 2021 for use in individuals aged ≥18 years.^2^ While Ad26.COV2.S (Johnson & Johnson’s Janssen) vaccine was also authorized by Health Canada in March 2021 for use in individuals aged ≥18 years, it has not been used widely in Canada.^3^ Very rare cases of thrombosis with thrombocytopenia have been reported globally following receipt of adenovirus vector vaccines.^4^ Rare adverse events such as myocarditis and pericarditis have been reported after receiving the mRNA vaccines in Canada and internationally, particularly among adolescent males and young adults.^5-13^ The observed number of myocarditis/pericarditis cases was higher than the expected number of cases among individuals aged 12–39 years in the US and Canada.^14-16^ Facial palsy, including Bell’s palsy has also been reported after the receipt of an mRNA COVID-19 vaccine, predominantly in adults aged 18–44 years and rarely in children aged ≤11 years.^17^ Rare cases of Guillain-Barré Syndrome (GBS) have been reported following vaccinations with ChAdOx1 and Ad26.COV2.S vaccines.^18-20^

On 26 October 2021, the US Food and Drug Administration recommended emergency use authorization for a two-dose primary series of BNT162b2 using one-third (10-μg) of the teen and adult dose in children aged 5–11 years.^21^ The US Centers for Disease Control and Prevention recommended this vaccine for children aged 5–11 years on 2 November 2021.^22^ On 19 November 2021, Health Canada authorized the BNT162b2 pediatric vaccine for use in children aged 5–11 years and on the same day Canada’s National Advisory Committee on Immunization issued their statement recommending its use.^23 24^ The phase 2/3 clinical trial of BNT162b2 in children aged 5-11 years observed no serious adverse events attributed to the vaccine, including myocarditis/pericarditis. However, only 1,518 vaccinated children and 750 children in the placebo group contributed to the safety data with at least 2 months of follow-up after receipt of the second dose; an additional 1,509 vaccinated children and 788 children in the placebo group with only 2.4 weeks of median follow-up after receipt of the second dose contributed to supplemental safety data. Thus, post-licensure surveillance is critical to identify vaccine safety signals for myocarditis, pericarditis, and other rare adverse events of special interest (AESI) relevant to this population.^25^

Background rates facilitate the identification of vaccine safety signals by permitting the calculation of expected numbers of events, which can be compared to observed events.^26-29^ Background rates may vary by calendar time, age, sex, socioeconomic status, and geography.^29^ There is a lack of data on recent background rates of potential AESI to inform COVID-19 vaccine safety surveillance in Ontario. We previously reported the background incidence rates of selected thromboembolic and coagulation disorders in Ontario.^30^ In this paper we report the background incidence rates of hospitalizations and emergency department visits for nine additional AESI during five pre-pandemic years (2015–2019) and 2020 in Ontario, Canada. The rates in children are presented according to vaccine eligible age bands to facilitate safety signal assessment in the pediatric population, specifically in children aged <12 years.

## Methods

We conducted a population-based retrospective observational study using linked health administrative databases from Ontario, Canada and analyzed at ICES (formerly the Institute for Clinical Evaluative Sciences). We identified hospitalizations and emergency department visits for 9 AESI: Bell’s palsy, idiopathic thrombocytopenia, febrile convulsions, acute disseminated encephalomyelitis, myocarditis, pericarditis, Kawasaki disease, Guillain-Barré syndrome, and transverse myelitis. Cases were identified using diagnostic codes from the *International Statistical Classification of Diseases and Related Health Problems, Tenth Revision, Canada (ICD-10-CA)*^31^ (Supplemental Table S1). Where available, we used codes that have been validated and/or used in previous studies.^26 32 33^ Hospitalizations and emergency department visits were identified from the Canadian Institute for Health Information (CIHI) Discharge Abstract Database (DAD) and CIHI’s National Ambulatory Care Reporting System, respectively. For hospitalizations, we included cases where an ICD-10-CA code of interest was present on admission and of primary relevance to the stay, and not listed as a secondary diagnosis, comorbidity, or as part of the medical history. We included emergency department visits where a code of interest was present in any diagnosis field. As Bell’s palsy cases are often treated in outpatient settings, we also included outpatient Bell’s palsy cases identified from the Ontario Health Insurance Plan physician billing claims database, and included these in a broader case definition combining hospitalizations, emergency department visits, and outpatient visits. For idiopathic thrombocytopenia, acute disseminated encephalomyelitis, and transverse myelitis, we also used narrow definitions to estimate conservative rates for these conditions (Supplemental Table S1). We included only new cases after a 365-day period without the same condition (i.e., group of codes included in each AESI definition) for each individual. Thus, one episode per individual was included across all data sources; multiple episodes of an AESI for an individual within a 365-day period were not included. We obtained information on age and sex from Ontario’s Registered Persons Database, which contains all Ontarians registered under the Ontario Health Insurance Plan.

### Statistical analysis

For each AESI, we calculated annual incidence rates per 100,000 population by age group (0–4, 5–11 years, 12–15, 16–19, 20–24, 25–29, 30–39, 40–49, 50–59, 60–69, 70–79 and ≥80 years), by sex, and by age group and sex during each of five pre-pandemic years (2015–2019) and 2020. We also estimated the overall mean annual incidence for 2015–2019. The rates of idiopathic thrombocytopenia using the narrow definition are presented for 0–29 years only because the rates for ≥30 years have been reported previously.^30^ We calculated monthly average rates for 2015–2019 to examine seasonality in children aged 0–11 years. We used Statistics Canada Census and intercensal population estimates as denominators. The 95% confidence intervals (CIs) were calculated using a gamma distribution.^34^

## Results

### Rates across all ages

The study population included approximately 85 million person-years of observation for all age groups from 2015 to 2020. The average annual study population was 14 million with 51% female.

During 2015-2019, the overall mean incidence rate of hospitalizations and ED visits for both sexes and all ages was highest for idiopathic thrombocytopenia (43.9 per 100,000) and lowest for Kawasaki disease (1.6 per 100,000). The mean incidence rates per 100,000 population for other AESI were 27.8 for Bell’s palsy, 25.0 for febrile convulsions, 22.8 for acute disseminated encephalomyelitis, 11.3 for myocarditis/pericarditis, 8.6 for pericarditis, 2.9 for myocarditis, 1.9 for Guillain-Barré syndrome, and 1.7 for transverse myelitis. The mean rate of Bell’s palsy was 71.3 per 100,000 population when cases identified in outpatient physician offices were included. Rates were generally consistent over time (Table 1). Rates were lower in 2020 than during the pre-pandemic years for febrile convulsions, acute disseminated encephalomyelitis, and Guillain-Barré syndrome. The annual rates of myocarditis/pericarditis, myocarditis, and pericarditis across all age groups were relatively stable during 2017–2019, and slightly lower in 2020 (Supplemental Table S6). Rates of pericarditis were approximately 3 times higher than rates of myocarditis.

**Table 1:** Annual background rates of adverse events of special interest (AESI) in Ontario from 2015 to 2020.

| Year | Incidence rate per 100,000 population (95% confidence interval) |  |  |  |  |  |  |
| --- | --- | --- | --- | --- | --- | --- | --- |
|  | Bell's palsy | Bell's palsy<br>(including<br>outpatient visits) | ITP, narrow<br>definition | ITP, broad<br>definition | Febrile<br>convulsions | ADEM, narrow<br>definition | ADEM, broad<br>definition |
| 2015 | 27.0<br>(26.1, 27.9) | 71.3<br>(69.9, 72.8) | 6.41<br>(6.00, 6.85) | 40.5<br>(39.4, 41.5) | 23.5<br>(22.7, 24.3) | 0.16<br>(0.10, 0.24) | 21.8<br>(21.1, 22.6) |
| 2016 | 27.0<br>(26.1, 27.9) | 71.7<br>(70.3, 73.1) | 6.31<br>(5.90, 6.74) | 43.5<br>(42.4, 44.6) | 28.57<br>(27.7, 29.5) | 0.22<br>(0.15, 0.31) | 23.0<br>(22.2, 23.8) |
| 2017 | 27.9<br>(27.0, 28.7) | 72.0<br>(70.6, 73.4) | 6.14<br>(5.74, 6.56) | 44.8<br>(43.7, 45.9) | 23.7<br>(22.9, 24.5) | 0.22<br>(0.15, 0.31) | 23.2<br>(22.4, 24.0) |
| 2018 | 27.9<br>(27.0, 28.7) | 70.9<br>(69.6, 72.3) | 5.75<br>(5.37, 6.16) | 44.9<br>(43.8, 46.0) | 25.5<br>(24.6, 26.3) | 0.20<br>(0.13, 0.28) | 23.0<br>(22.2, 23.8) |
| 2019 | 29.2<br>(28.3, 30.1) | 70.6<br>(69.2, 72.0) | 5.81<br>(5.42, 6.21) | 45.8<br>(44.8, 47.0) | 24.0<br>(23.2, 24.8) | 0.14<br>(0.09, 0.22) | 22.9<br>(22.1, 23.7) |
| 2020 | 31.4<br>(30.5, 32.3) | 70.5<br>(69.1, 71.9) | 4.54<br>(4.20, 4.90) | 46.5<br>(45.4, 47.6) | 13.5<br>(12.9, 14.1) | 0.10<br>(0.06, 0.17) | 20.1<br>(19.4, 20.8) |
| Year | Myocarditis/peri<br>carditis | Myocarditis | Pericarditis | Kawasaki disease | Guillain-Barré<br>syndrome | Transverse<br>myelitis, narrow<br>definition | Transverse<br>myelitis, broad<br>definition |
| 2015 | 9.82<br>(9.30, 10.36) | 2.54<br>(2.28, 2.82) | 7.53<br>(7.08, 8.00) | 1.96<br>(1.73, 2.21) | 1.83<br>(1.61, 2.07) | 0.61<br>(0.49, 0.76) | 1.27<br>(1.09, 1.47) |
| 2016 | 10.22<br>(9.69, 10.77) | 2.88<br>(2.60, 3.17) | 7.64<br>(7.19, 8.11) | 2.00<br>(1.77, 2.25) | 1.94<br>(1.71, 2.18) | 0.90<br>(0.75, 1.07) | 1.84<br>(1.63, 2.09) |
| 2017 | 12.4<br>(11.8, 13.0) | 3.38<br>(3.08, 3.69) | 9.35<br>(8.85, 9.87) | 2.03<br>(1.80, 2.27) | 1.83<br>(1.61, 2.06) | 0.73<br>(0.60, 0.89) | 1.69<br>(1.48, 1.92) |
| 2018 | 11.9<br>(11.3, 12.4) | 2.58<br>(2.32, 2.86) | 9.44<br>(8.94, 9.96) | 2.15<br>(1.92, 2.41) | 1.88<br>(1.66, 2.12) | 0.85<br>(0.71, 1.02) | 1.81<br>(1.60, 2.04) |
| 2019 | 12.0<br>(11.4, 12.6) | 2.93<br>(2.66, 3.22) | 9.24<br>(8.75, 9.75) | 1.71<br>(1.51, 1.94) | 1.82<br>(1.60, 2.05) | 0.87<br>(0.73, 1.04) | 1.95<br>(1.73, 2.19) |
| 2020 | 10.7<br>(10.2, 11.2) | 2.42<br>(2.18, 2.69) | 8.44<br>(7.97, 8.92) | 1.55<br>(1.35, 1.76) | 1.34<br>(1.16, 1.54) | 0.83<br>(0.69, 1.00) | 1.93<br>(1.71, 2.17) |

ADEM = Acute disseminated encephalomyelitis, ITP = Idiopathic thrombocytopenia

Mean pre-pandemic incidence rates of acute disseminated encephalomyelitis and transverse myelitis tended to be higher for females than males (Figure 1). In contrast, mean pre-pandemic rates of myocarditis, pericarditis, and Guillain-Barré syndrome were higher for males than females. Rates for Bell’s palsy, acute disseminated encephalomyelitis, myocarditis, pericarditis, and Guillain-Barré syndrome generally increased with age. Mean pre-pandemic rates of idiopathic thrombocytopenia increased with age after 24 years of age, and was higher in males than females at ≥40 years. Mean pre-pandemic rates of myocarditis/pericarditis were similar for females and males up to 11 years of age, then diverged to be higher for males than females, with two peaks in males at 16–24 years and ≥70 years (Figure 1). The mean rates of myocarditis were higher for males than females aged 12–59 years; rates of pericarditis were higher for males aged ≥12 years. Febrile convulsions and Kawasaki disease were predominantly childhood diseases and generally decreased with age.

**Figure 1:**
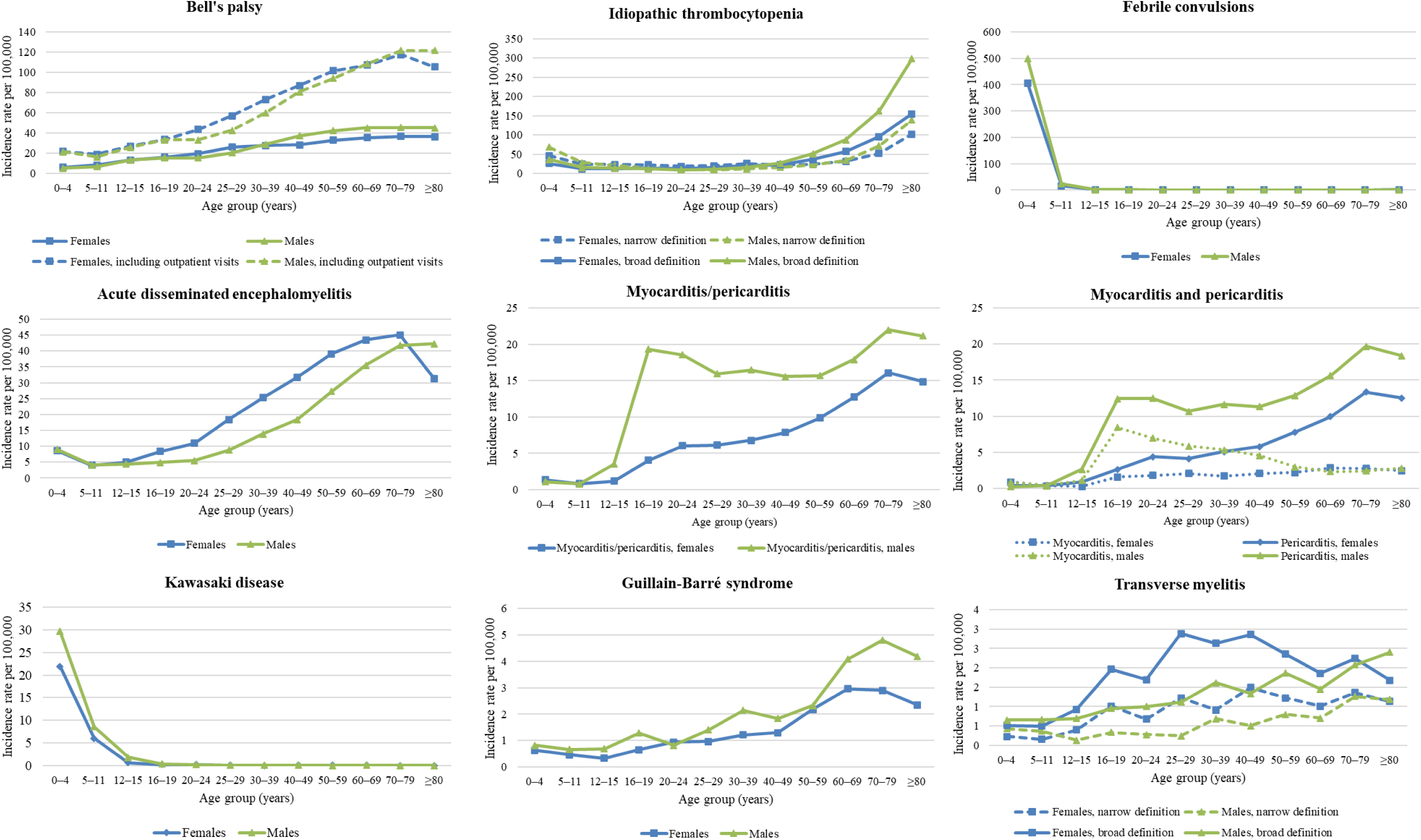
Age and sex stratified mean annual background rates (per 100,000 population) for nine adverse events of special interest in Ontario, 2015 to 2019

### Rates in children aged 0–11 years

The study population included approximately 10.7 million person-years of observation among children aged 0–11 years from 2015 to 2020; 60% of the study population was aged 5–11 years. The average annual study population was 1.8 million, with 49% female.

During 2015–2019, the mean rate of hospitalizations and ED visits was highest for febrile convulsion (453 per 100,000 in children aged 0–4 years and 20.1 per 100,000 in children aged 5– 11 years) and lowest for pericarditis (0.4 per 100,000 in both age groups). For children aged 0–4 years, the mean rates per 100,000 population for other AESI were 31.3 for idiopathic thrombocytopenia, 25.9 for Kawasaki disease, 8.8 for acute disseminated encephalomyelitis, 5.5 for Bell’s palsy, 1.2 for myocarditis/pericarditis, 0.9 for myocarditis, 0.7 for Guillain-Barré syndrome, and 0.6 for transverse myelitis; the rates for children aged 5–11 years were 13.6 for idiopathic thrombocytopenia, 7.4 for Bell’s palsy, 7.3 for Kawasaki disease, 4.1 for acute disseminated encephalomyelitis, 0.8 for myocarditis/pericarditis, and 0.6 for transverse myelitis and Guillain-Barré syndrome, 0.5 for myocarditis. When cases identified in outpatient physician offices were added, the mean rates per 100,000 population for Bell’s palsy was 21.3 among children aged 0–4 years, and 17.4 among children aged 5–11 years.

The annual rates for most of the studied AESI fluctuated over the study period and were lower in 2020 than the mean annual rate during the pre-pandemic years (2015–2019) except for Guillain-Barré syndrome among children aged 0–4 years and pericarditis among children aged 5–11 years (Supplemental Tables S2-S9). Rates of Kawasaki disease among children aged 0–4 years were 3–4 times higher than children aged 5–11 years.

Rates of idiopathic thrombocytopenia, febrile convulsions, and Kawasaki disease generally tended to be higher for males than females for both age groups (Supplemental Tables S3, S4, S7). In contrast, rates of hospitalizations, emergency department visits, and outpatient visits for Bell’s palsy tended to be higher for females than males in children aged 5–11 years (Supplemental Table S2).

We noted seasonality for three AESI: Kawasaki disease peaked during November through March for children aged 0–4 years and during December and January for children aged 5–11 years; febrile convulsions peaked during December through March for children aged 0–4 years; and idiopathic thrombocytopenia among children aged 0–4 years peaked in December, January, and April when using the narrow definition and in December and January when using the broad definition (Figure 2).

**Figure 2:**
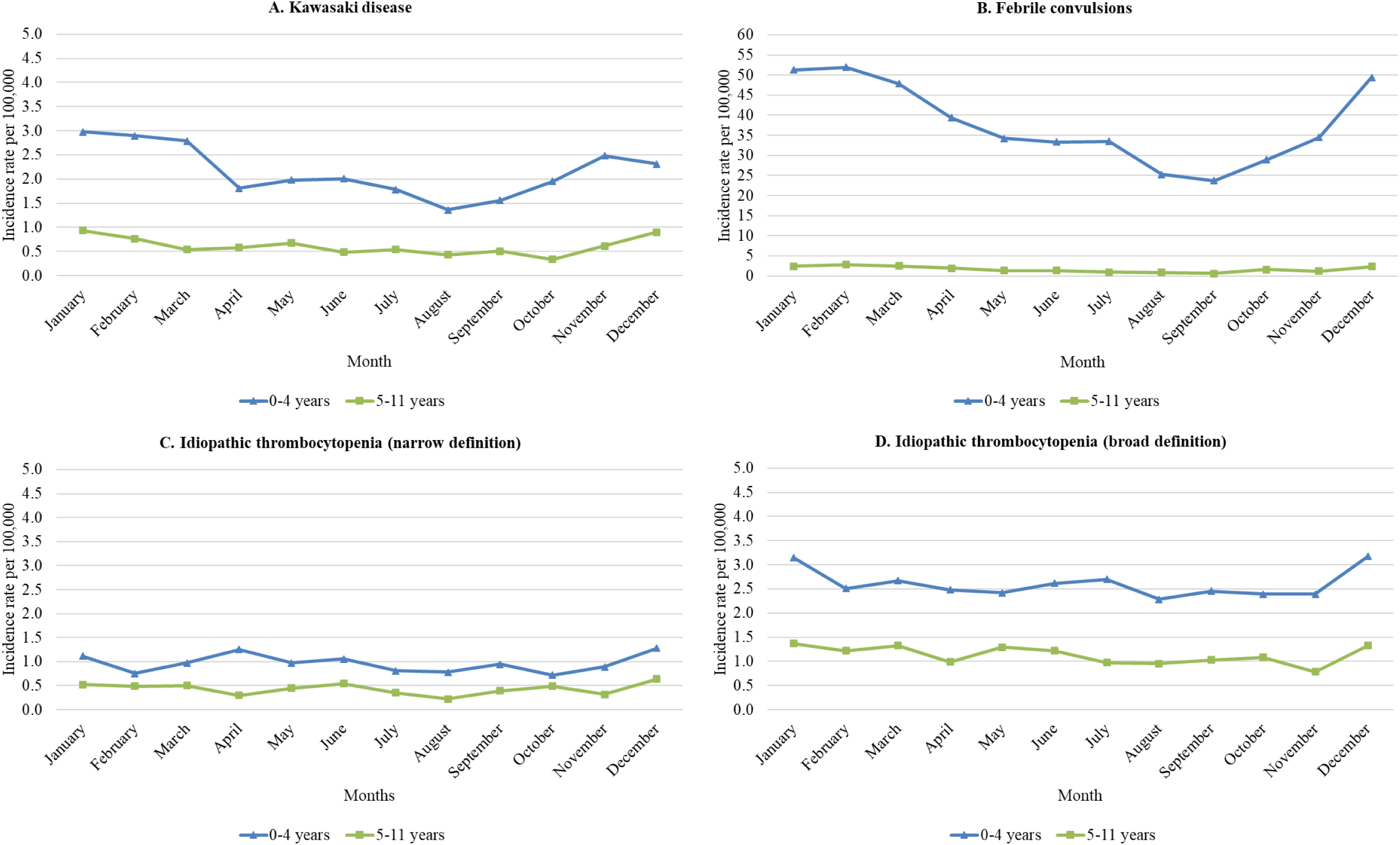
Monthly average incidence rates of hospitalization and emergency department visits in Ontario, 2015 to 2020: A. Kawasaki disease, B. Febrile convulsions, C. Idiopathic thrombocytopenia (narrow definition), D. Idiopathic thrombocytopenia (broad definition).

## Discussion

In this study, we estimated background rates of hospitalizations and emergency department visits for selected AESI in during 2015 to 2020 in Ontario. Our findings suggest that overall rates were generally consistent over time. Females had higher rates of acute disseminated encephalomyelitis and transverse myelitis while males had higher rates of myocarditis, pericarditis, and Guillain-Barré syndrome. Bell’s palsy, acute disseminated encephalomyelitis, and Guillain-Barré syndrome increased with age. Rates of pericarditis were higher than myocarditis. The rates of myocarditis and/or pericarditis increased with age until 79 years; and the rates were higher in males from 12–59 years for myocarditis and in males aged ≥12 years, with two peaks for pericarditis. However, in children aged <12 years, rates varied during the pre-pandemic period and were lower in 2020 for most of these AESI. Males aged 0–11 years had higher rates of febrile convulsions, Kawasaki disease, and idiopathic thrombocytopenia than females aged 0–11 years, whereas females aged 5–11 years had higher rates of Bell’s palsy than their male counterparts. Kawasaki disease and febrile convulsions tended to peak during the winter months.

We observed increased rates of Bell’s palsy, Guillain-Barré syndrome, idiopathic thrombocytopenia and myocarditis and/or pericarditis with age similar to previous studies.^35^ Contrary to our finding, a gradual decrease in incidence of myocarditis with age in adults was reported in a previous study in Finland.^36^ We observed a higher rate in older children aged 16–19 years than younger children similar to other studies conducted in hospitalized children.^37^ Similar to our study, rates of myocarditis or pericarditis have been observed to increase with age with higher rates in males in a multinational study.^35^

Among children aged <12 years, compared to the mean rates during the pre-pandemic years, we observed lower rates in 2020 for most of these AESI except for Guillain-Barré syndrome in children aged 0–4 years, and pericarditis in children aged 5–11 years. The most striking absolute reduction in rate was observed for febrile convulsions in children aged 0–4 years. This likely resulted from the decreased circulation of non-COVID-19 respiratory viruses during the pandemic^38^ as a result of broad public health measures in Ontario, including the lockdown in March-June 2020, continued province-wide mask mandate, and school-based strategies to ensure safe reopening of schools for in-person attendance in fall 2020.^39^ A decrease in healthcare-seeking behaviours in 2020, along with surpassed hospital capacity at times of peak COVID-19 circulation, may also have caused some reduction in rates of hospital admissions or emergency department visits.

As expected, rates of Bell’s palsy were higher when outpatient cases were added to cases that were hospitalized or sought care in emergency departments. However, the diagnostic code used to identify outpatient Bell’s palsy cases from the physician billing claims also includes the diagnosis facial palsy. Consequently, the rates including outpatient cases may be an overestimate of the rate of Bell’s palsy. The rate of Bell’s palsy including outpatient and inpatient cases in US children aged 0–17 years was reported to be 24 per 100,000 person-years.^26^ This rate is comparable to our estimated pre-pandemic mean rate for inpatient, ED visit, and outpatient cases for children aged 0–4 years (21.3 per 100,000) but higher than our rate for children aged 5–11 years (17.4 per 100,000). The background rate of Bell’s palsy was reported to be 15 per 100,000 person-years in both males and females aged 1–5 years, and 25 and 21 per 100,000 person-years in females and males aged 6–17 years, respectively, in a recent multinational study using electronic health records and health claims data, including primary care data.^35^ The differences in the background rates between our study and other studies likely resulted from differences in calendar time, geography, population characteristics, distribution of risk factors, local transmission of possible causative viral infections, and healthcare systems.

Our estimated background rates of Kawasaki disease are within the previously reported range of 19.1–32.1 per 100,000 in Canadian children younger than 5 years of age^32^ and 5.1–50.4 per 100,000 in US children aged 0–6 years of age.^40^ Similar to those studies, we also observed higher rates for males than females among children aged 0–4 years. Our estimated pre-pandemic mean rates of GBS are similar to the rates reported in children aged 1–9 years in Denmark.^41^ Our mean rates of transverse myelitis are higher than the rates reported in children aged 1–9 years in Denmark (0.36 per 100,000 person-years for children aged 4–9 years) and Israel (0.40 per 100,000 person-years in children aged 0–9 years).^26 41^ We observed seasonality for Kawasaki disease and febrile convulsions in young children with peak rates during the winter months conforming to previous reports.^42 43^

There are some limitations of this study. Our rates may be higher than those reported in the literature using hospitalization data alone as we also included cases treated in emergency departments. Imperfect validity of the diagnostic codes in administrative data without information on clinical and/or diagnostic confirmation may have resulted in under or overestimation. However, we used previously validated codes and/or codes that have been used in previous studies^26 32 33^ to improve the accuracy of case ascertainment in administrative data. The quality of DAD hospitalization data has been previously evaluated.^44^ Our estimated rates may not be generalizable to other populations or settings because background rates are population-specific and differ by calendar time, population structure, distribution of risk factors, and healthcare systems.^29^

Our estimated background rates of hospitalizations and emergency department visits for the selected AESI in all age groups will facilitate estimating the number of expected events for these conditions. Reports of these AESI arising from Ontario’s passive vaccine safety surveillance data can be compared to determine if they are greater than the expected number of events to assess potential safety signals. Additionally, our estimates suggest that some of these AESI are common in children aged <12 years and some have demonstrated seasonality. This information will further aid clinicians and public health authorities to gauge and contextualize higher observed events following immunization for these AESI.

## Data Availability

The dataset from this study is held securely in coded form at ICES. While legal data sharing agreements between ICES and data providers (e.g., healthcare organizations and government) prohibit ICES from making the dataset publicly available, access may be granted to those who meet pre-specified criteria for confidential access, available at www.ices.on.ca/DAS. The full dataset creation plan and underlying analytic code are available from the authors upon request, understanding that the computer programs may rely upon coding templates or macros that are unique to ICES and are therefore either inaccessible or may require modification.

## Contributors

JCK, SN, NT, SEW conceived of the study design. JCK oversaw the study. AC prepared data and performed the statistical analysis. SN drafted the manuscript. All authors interpreted the results, critically revised the manuscript, and approved the final version.

## Ethics approval

ICES is a prescribed entity under Ontario’s Personal Health Information Protection Act (PHIPA). Section 45 of PHIPA authorizes ICES to collect personal health information, without consent, for the purpose of analysis or compiling statistical information with respect to the management of, evaluation or monitoring of, the allocation of resources to or planning for all or part of the health system. Projects that use data collected by ICES under section 45 of PHIPA, and use no other data, are exempt from REB review. The use of the data in this project is authorized under section 45 and approved by ICES’ Privacy and Legal Office.

