## Supplemental Table for "Background incidence rates of adverse events of special interest related to COVID-19 vaccines in Ontario, Canada, 2015 to 2020, to inform COVID-19 vaccine safety surveillance"

Supplemental Table S1: ICD-10-CA diagnostic codes

| AESI | Definitions | Diagnosis/ICD-10 label | ICD-10-CA code |
|---|---|---|---|
| Bell’ s palsy | Bell’s palsy | Bell’s palsy | G51.0 |
| | Bell’s palsy, including outpatient visits | Bell’s palsy | G51.0 |
| | | Bell’s palsy, facial nerve disorders | OHIP code: 351 |
| Idiopathic thrombocytopenia | Idiopathic thrombocytopenia, narrow definition | Idiopathic thrombocytopenic purpura | D69.3x |
| | Idiopathic thrombocytopenia, broad definition | Idiopathic thrombocytopenic purpura | D69.3x |
| | | Other primary thrombocytopenia | D69.4 |
| | | Secondary thrombocytopenia | D69.5 |
| | | Thrombocytopenia, unspecified | D69.6 |
| | | Wiskott-Aldrich syndrome | D82.0 |
| | | Thrombotic microangiopathy<br>Includes: Thrombotic thrombocytopenic purpura | M31.1 |
| Febrile convulsions | Febrile convulsions | Febrile convulsions | R56.0x |
| Acute disseminated encephalomyelitis | Acute disseminated encephalomyelitis, narrow definition | Acute disseminated encephalomyelitis | G04.0 |
| | Acute disseminated encephalomyelitis, broad definition | Encephalitis, myelitis, and encephalomyelitis | G04.x |
| | | Encephalitis, myelitis, and encephalomyelitis in diseases classified elsewhere | G05.x |
| | | Multiple sclerosis | G35 |
| | | Other acute disseminated demyelination | G36.x |
| | | Other demyelinating diseases of central nervous system | G37.x |
| | | Toxic encephalopathy | G92 |
| | | Encephalopathy, unspecified | G93.4 |
| | | Disorder of central nervous system, unspecified | G96.9 |
| Myocarditis/pericarditis | Myocarditis/pericarditis | Acute pericarditis | I30.x |
| | | Pericarditis in disease classified elsewhere | I32.x |
| | | Acute myocarditis | I40.x |
| | | Myocarditis in diseases classified elsewhere | I41.x |
| | | Myocarditis, unspecified | I51.4 |
| | Myocarditis | Acute myocarditis | I40.x |
| | | Myocarditis in diseases classified elsewhere | I41.x |
| | | Myocarditis, unspecified | I51.4 |
| | Pericarditis | Acute pericarditis | I30.x |
| | | Pericarditis in disease classified elsewhere | I32.x |
| Kawasaki disease | Kawasaki disease | Mucocutaneous lymph node syndrome (Kawasaki) | M30.3 |

| | | | |
|---|---|---|---|
| Guillain-Barré syndrome | Guillain-Barré syndrome | Guillain-Barré syndrome | G61.0 |
| Transverse myelitis | Transverse myelitis, narrow definition | Acute transverse myelitis in demyelinating disease of central nervous system | G37.3 |
| | Transverse myelitis, broad definition | Neuromyelitis optica [Devic] | G36.0 |
| | | Diffuse sclerosis | G37.0 |
| | | Acute transverse myelitis in demyelinating disease of central nervous system | G37.3 |
| | | Other specified demyelinating diseases of central nervous system | G37.8 |
| | | Demyelinating disease of central nervous system, unspecified | G37.9 |

Supplemental Table S2: Annual background rates of hospitalizations and emergency department visits for Bell's palsy by age group and sex in Ontario, 2015 to 2020

| Age group, years | Incidence rate per 100,000 population (95% confidence interval) | | | | | |
|---|---|---|---|---|---|---|
| | **2015** | **2016** | **2017** | **2018** | **2019** | **2020** |
| **Bell's palsy** | | | | | | |
| **Both sexes** | | | | | | |
| All ages | 27.0 (26.1, 27.9) | 27.0 (26.1, 27.9) | 27.9 (27.0, 28.7) | 27.9 (27.0, 28.7) | 29.2 (28.3, 30.1) | 31.4 (30.5, 32.3) |
| 0–4 | 6.59 (4.84, 8.76) | 6.13 (4.45, 8.22) | 5.58 (3.99, 7.60) | 5.01 (3.51, 6.93) | 4.30 (2.92, 6.11) | 2.63 (1.58, 4.10) |
| 5–11 | 5.89 (4.51, 7.55) | 8.09 (6.47, 9.99) | 8.14 (6.52, 10.0) | 7.24 (5.73, 9.04) | 7.59 (6.04, 9.42) | 5.84 (4.49, 7.47) |
| 12–15 | 13.5 (10.8, 16.8) | 12.8 (10.2, 16.0) | 13.8 (11.0, 17.0) | 13.5 (10.8, 16.7) | 12.1 (9.5, 15.1) | 14.5 (11.7, 17.8) |
| 16–19 | 17.0 (14.1, 20.4) | 14.5 (11.8, 17.7) | 15.6 (12.8, 18.9) | 16.1 (13.3, 19.4) | 15.1 (12.4, 18.3) | 16.9 (14.0, 20.2) |
| 20–24 | 15.5 (13.1, 18.3) | 17.9 (15.3, 20.8) | 18.4 (15.8, 21.3) | 16.6 (14.2, 19.3) | 17.7 (15.2, 20.5) | 19.8 (17.2, 22.7) |
| 25–29 | 21.3 (18.4, 24.5) | 22.8 (19.9, 26.1) | 23.2 (20.2, 26.4) | 24.5 (21.6, 27.8) | 23.5 (20.6, 26.6) | 22.5 (19.7, 25.5) |
| 30–39 | 27.8 (25.5, 30.4) | 28.9 (26.5, 31.5) | 26.7 (24.4, 29.1) | 28.1 (25.8, 30.6) | 29.1 (26.7, 31.5) | 30.3 (27.9, 32.8) |
| 40–49 | 33.3 (30.7, 36.0) | 30.2 (27.8, 32. 8) | 33.9 (31.3, 36.6) | 32.0 (29.5, 34.7) | 33.9 (31.3, 36.7) | 37.6 (34.9, 40.5) |
| 50–59 | 34.5 (32.0, 37.1) | 38.1 (35.5, 40.9) | 38.4 (35.7, 41.1) | 35.9 (33.4, 38.6) | 40.6 (37.9, 43.4) | 44.9 (42.1, 47.9) |
| 60–69 | 42.4 (39.2, 45.8) | 39.3 (36.3, 42.5) | 37.3 (34.4, 40.4) | 40.1 (37.1, 43.2) | 40.9 (38.0, 44.1) | 43.0 (40.0, 46.2) |
| 70–79 | 37.9 (34.0, 42.2) | 35.6 (31.8, 39.6) | 44.0 (39.9, 48.3) | 42.8 (38.9, 46.9) | 43.4 (39.6, 47.5) | 49.4 (45.4, 53.6) |
| ≥80 | 36.0 (31.3, 41.3) | 34.5 (30.0, 39.6) | 37.1 (32.4, 42.3) | 43.4 (38.4, 48.9) | 47.7 (42.5, 53.4) | 51.5 (46.2, 57.3) |
| **Females** | | | | | | |
| All ages | 25.3 (24.2, 26.6) | 24.9 (23.7, 26.1) | 26.8 (25.7, 28.1) | 26.3 (25.1, 27.5) | 27.4 (26.2, 28.6) | 29.5 (28.3, 30.8) |
| 0–4 | 6.60 (4.18, 9.90) | 6.55 (4.15, 9.83) | 6.57 (4.17, 9.86) | 4.28 (2.40, 7.06) | 5.70 (3.48, 8.80) | 3.12 (1.56, 5.59) |
| 5–11 | 6.77 (4.72, 9.42) | 7.86 (5.64, 10.7) | 8.57 (6.25, 11.5) | 8.90 (6.54, 11.8) | 9.06 (6.68, 12.0) | 7.95 (5.73, 10.7) |
| 12–15 | 12.7 (9.00, 17.5) | 12.6 (8.93, 17.3) | 13.6 (9.73, 18.4) | 13.1 (9.39, 17.9) | 14.3 (10.4, 19.2) | 14.1 (10.3, 18.9) |
| 16–19 | 17.9 (13.6, 23.0) | 15.1 (11.2, 19.9) | 17.3 (13.2, 22.4) | 15.9 (11.9, 20.7) | 14.1 (10.4, 18.7) | 14.8 (11.0, 19.5) |
| 20–24 | 18.5 (14.8, 22.9) | 19.7 (15.8, 24.2) | 20.6 (16.7, 25.1) | 19.6 (15.8, 23.9) | 17.9 (14.4, 22.0) | 18.7 (15.1, 23.0) |
| 25–29 | 22.3 (18.2, 27.1) | 26.2 (21.7, 31.3) | 26.8 (22.3, 31.9) | 28.9 (24.3, 34.1) | 26.2 (21.9, 31.0) | 22.6 (18.7, 27.1) |
| 30–39 | 28.5 (25.1, 32.2) | 26.6 (23.3, 30.1) | 28.0 (24.7, 31.6) | 26.1 (23.0, 29.5) | 28.1 (24.9, 31.6) | 30.3 (27.0, 33.9) |
| 40–49 | 27.4 (24.1, 30.9) | 25.5 (22.4, 28.9) | 30.5 (27.1, 34.3) | 28.2 (25.0, 31.8) | 29.2 (25.9, 32.9) | 32.0 (28.6, 35.9) |
| 50–59 | 30.4 (27.1, 33.9) | 34.5 (31.0, 38.2) | 33.2 (29.8, 36.8) | 30.4 (27.1, 33.9) | 35.1 (31.5, 38.8) | 39.9 (36.1, 43.9) |
| 60–69 | 36.1 (32.1, 40.6) | 33.2 (29.4, 37.4) | 34.7 (30.9, 39.0) | 36.3 (32.4, 40.5) | 36.3 (32.4, 40.4) | 37.8 (34.0, 42.1) |
| 70–79 | 36.5 (31.3, 42.3) | 31.7 (26.9, 37.0) | 38.5 (33.4, 44.2) | 38.4 (33.4, 43.9) | 38.4 (33.5, 43.8) | 44.7 (39.6, 50.4) |
| ≥80 | 33.1 (27.4, 39.7) | 29.1 (23.8, 35.2) | 34.5 (28.7, 41.0) | 39.9 (33.8, 46.8) | 44.7 (38.3, 51.9) | 52.3 (45.4, 60.0) |
| **Males** | | | | | | |

| | | | | | | |
|---|---|---|---|---|---|---|
| All ages | 28.6 (27.4, 30.0) | 29.2 (27.9, 30.5) | 28.9 (27.6, 30.2) | 29.5 (28.2, 30.8) | 31.0 (29.7, 32.3) | 33.3 (32.0, 34.7) |
| 0–4 | 6.58 (4.21, 9.79) | 5.72 (3.54, 8.74) | 4.63 (2.70, 7.41) | 5.69 (3.52, 8.70) | 2.97 (1.48, 5.32) | 2.16 (0.93, 4.25) |
| 5–11 | 5.03 (3.32, 7.32) | 8.32 (6.07, 11.1) | 7.72 (5.56, 10.4) | 5.65 (3.84, 8.02) | 6.18 (4.28, 8.63) | 3.82 (2.36, 5.84) |
| 12–15 | 14.3 (10.4, 19.2) | 13.0 (9.35, 17.7) | 14.0 (10.2, 18.7) | 13.9 (10.1, 18.6) | 9.98 (6.83, 14.1) | 14.8 (10.9, 19.7) |
| 16–19 | 16.2 (12.3, 21.0) | 13.9 (10.3, 18.4) | 14.0 (10.4, 18.5) | 16.3 (12.4, 21.0) | 16.1 (12.2, 20.8) | 18.8 (14.6, 23.93 |
| 20–24 | 12.8 (9.8, 16.4) | 16.3 (12.9, 20.2) | 16.5 (13.2, 20.4) | 13.9 (10.9, 17.4) | 17.6 (14.2, 21.5) | 20.8 (17.1, 25.0) |
| 25–29 | 20.2 (16.4, 24.7) | 19.7 (15.9, 24.0) | 19.7 (16.0, 24.0) | 20.4 (16.7, 24.7) | 20.9 (17.3, 25.2) | 22.3 (18.6, 26.6) |
| 30–39 | 27.2 (23.8, 30.8) | 31.4 (27.8, 35.2) | 25.3 (22.2, 28.8) | 30.1 (26.7, 33.8) | 30.1 (26.7, 33.7) | 30.3 (27.0, 33.9) |
| 40–49 | 39.4 (35.4, 43.6) | 35.1 (31.3, 39.1) | 37.4 (33.5, 41.6) | 36.0 (32.2, 40.2) | 38.9 (34.9, 43.4) | 43.5 (39.3, 48.0) |
| 50–59 | 38.6 (34.9, 42.6) | 41.8 (38.0, 46.0) | 43.6 (39.7, 47.9) | 41.5 (37.7, 45.7) | 46.2 (42.1, 50.6) | 50.0 (45.8, 54.6) |
| 60–69 | 49.1 (44.2, 54.4) | 45.9 (41.3, 51.0) | 40.1 (35.8, 44.8) | 44.2 (39.7, 49.0) | 46.0 (41.5, 50.8) | 48.5 (43.9, 53.4) |
| 70–79 | 39.5 (33.7, 46.1) | 40.1 (34.3, 46.5) | 50.2 (44.0, 57.1) | 47.8 (41.9, 54.4) | 49.2 (43.3, 55.7) | 54.6 (48.5, 61.3) |
| ≥80 | 40.6 (32.7, 49.8) | 43.0 (35.0, 52.2) | 41.1 (33.4, 50.1) | 48.7 (40.4, 58.1) | 52.2 (43.7, 61.8) | 50.4 (42.2, 59.8) |
| **Bell’s palsy (including outpatient visits)** | | | | | | |
| **Both sexes** | | | | | | |
| All ages | 71.3 (69.9, 72.8) | 71.7 (70.3, 73.1) | 72.0 (70.6, 73.4) | 70.9 (69.6, 72.3) | 70.6 (69.2, 72.0) | 70.5 (69.1, 71.9) |
| 0–4 | 23.1 (19.7, 26.9) | 22.0 (18.7, 25.7) | 22.0 (18.7, 25.8) | 22.1 (18.8, 25.8) | 17.5 (14.6, 20.8) | 17.0 (14.1, 20.3) |
| 5–11 | 15.4 (13.1, 17.9) | 16.6 (14.2, 19.2) | 19.6 (17.1, 22.5) | 18.4 (15.9, 21.1) | 16.9 (14.6, 19.6) | 14.6 (12.4, 17.0) |
| 12–15 | 26.4 (22.5, 30.8) | 26.0 (22.1, 30.3) | 30.5 (26.3, 35.1) | 23.3 (19.7, 27.5) | 24.5 (20.8, 28.7) | 28.0 (24.0, 32.4) |
| 16–19 | 34.0 (29.8, 38.7) | 34.6 (30.3, 39.3) | 34.1 (29.9, 38.8) | 32.8 (28.7, 37.3) | 31.2 (27.2, 35.7) | 31.0 (27.0, 35.4) |
| 20–24 | 38.2 (34.3, 42.3) | 38.9 (35.0, 43.0) | 38.8 (35.0, 42.9) | 37.4 (33.7, 41.4) | 37.1 (33.5, 41.0) | 38.3 (34.6, 42.2) |
| 25–29 | 49.7 (45.2, 54.4) | 48.8 (44.5, 53.5) | 52.5 (48.0, 57.3) | 49.5 (45.2, 54.0) | 46.7 (42.6, 51.0) | 48.3 (44.2, 52.6) |
| 30–39 | 68.8 (65.0, 72.8) | 68.0 (64.2, 71.8) | 67.3 (63.6, 71.1) | 64.8 (61.2, 68.5) | 64.6 (61.1, 68.2) | 64.0 (60.6, 67.6) |
| 40–49 | 85.5 (81.4, 89.8) | 83.9 (79.8, 88.2) | 84.8 (80.6, 89.1) | 84.0 (79.9, 88.3) | 81.0 (77.0, 85.2) | 83.3 (79.2, 87.5) |
| 50–59 | 94.9 (90.8, 99.2) | 102 (97.3, 106) | 99.0 (94.8, 103) | 93.4 (89.3, 97.7) | 101 (96.6, 105) | 99.4 (95.1, 104) |
| 60–69 | 111 (106, 116) | 113 (107, 118) | 103 (98.6, 108) | 109 (104, 114) | 103 (98.7, 108) | 105 (101, 110) |
| 70–79 | 121 (113, 128) | 113 (106, 120) | 122 (116, 129) | 121 (115, 128) | 120 (114, 127) | 114 (108, 121) |
| ≥80 | 108 (99.4, 117) | 104 (96.2, 113) | 108 (100, 117) | 117 (109, 126) | 121 (112, 130) | 116 (108, 124) |
| **Females** | | | | | | |
| All ages | 74.3 (72.3, 76.4) | 74.7 (72.7, 76.7) | 76.3 (74.2, 78.3) | 74.3 (72.3, 76.3) | 74.2 (72.2, 76.2) | 73.9 (72.0, 75.9) |
| 0–4 | 21.5 (16.9, 27.0) | 19.1 (14.8, 24.2) | 24.0 (19.2, 29.7) | 23.1 (18.4, 28.7) | 19.4 (15.1, 24.6) | 17.3 (13.3, 22.3) |
| 5–11 | 16.5 (13.1, 20.3) | 16.7 (13.4, 20.6) | 21.1 (17.4, 25.5) | 20.3 (16.6, 24.5) | 18.7 (15.2, 22.8) | 16.3 (13.2, 20.3) |
| 12–15 | 26.1 (20.6, 32.6) | 26.2 (20.8, 32.7) | 32.8 (26.6, 39.9) | 23.7 (18.5, 29.8) | 25.3 (20.0, 31.6) | 28.9 (23.2, 35.5) |
| 16–19 | 32.7 (26.8, 39.5) | 35.1 (29.0, 42.1) | 35.6 (29.5, 42.6) | 34.7 (28.7, 41.6) | 29.9 (24.4, 36.3) | 30.2 (24.6, 36.6) |
| 20–24 | 45.0 (39.0, 51.6) | 43.1 (37.3, 49.6) | 44.7 (38.9, 51.2) | 44.5 (38.8, 50.9) | 39.6 (34.3, 45.6) | 36.9 (31.7, 42.6) |
| 25–29 | 54.5 (48.0, 61.8) | 55.8 (49.2, 63.0) | 62.7 (55.6, 70.2) | 58.4 (51.8, 65.6) | 52.4 (46.3, 59.0) | 52.7 (46.7, 59.4) |

| | | | | | | |
|---|---|---|---|---|---|---|
| 30–39 | 76.5 (70.9, 82.3) | 74.4 (69.0, 80.1) | 77.1 (71.6, 83.0) | 68.0 (62.9, 73.4) | 69.9 (64.8, 75.3) | 74.0 (68.8, 79.4) |
| 40–49 | 87.2 (81.4, 93.3) | 86.0 (80.2, 92.1) | 88.6 (82.8, 94.8) | 87.7 (81.8, 93.8) | 85.1 (79.3, 91.2) | 87.4 (81.6, 93.6) |
| 50–59 | 102 (96.1, 108) | 106 (99.6, 112) | 103 (97.1, 110) | 92.6 (86.9, 98.6) | 105 (98.8, 111) | 99.4 (93.4, 106) |
| 60–69 | 108 (101, 115) | 113 (106, 120) | 101 (94.3, 108) | 110 (103, 117) | 105 (98.0, 112) | 104 (97.7, 111) |
| 70–79 | 116 (106, 126) | 110 (101, 120) | 122 (113, 132) | 122 (113, 132) | 118 (109, 127) | 115 (106, 124) |
| ≥80 | 99.1 (89.0, 110) | 97.5 (87.6, 108) | 101 (91.2, 112) | 111 (101, 122) | 117 (107, 129) | 117 (107, 128) |
| **Males** | | | | | | |
| All ages | 68.3 (66.3, 70.3) | 68.7 (66.7, 70.7) | 67.5 (65.6, 69.5) | 67.5 (65.6, 69.5) | 66.9 (65.0, 68.8) | 67.0 (65.1, 68.9) |
| 0–4 | 24.7 (19.8, 30.3) | 24.8 (20.0, 30.4) | 20.2 (15.8, 25.3) | 21.2 (16.7, 26.4) | 15.7 (11.9, 20.3) | 16.7 (12.8, 21.4) |
| 5–11 | 14.4 (11.3, 17.9) | 16.5 (13.2, 20.3) | 18.2 (14.8, 22.2) | 16.6 (13.4, 20.4) | 15.3 (12.2, 18.9) | 12.7 (9.92, 16.1) |
| 12–15 | 26.7 (21.3, 33.1) | 25.8 (20.5, 32.0) | 28.2 (22.7, 34.8) | 23.0 (18.1, 29.0) | 23.7 (18.7, 29.7) | 27.2 (21.8, 33.5) |
| 16–19 | 35.3 (29.3, 42.0) | 34.1 (28.3, 40.7) | 32.8 (27.1, 39.3) | 31.0 (25.5, 37.3) | 32.5 (26.9, 38.9) | 31.8 (26.2, 38.2) |
| 20–24 | 31.9 (27.1, 37.3) | 35.0 (30.0, 40.6) | 33.4 (28.6, 38.8) | 30.9 (26.4, 36.1) | 34.8 (30.0, 40.1) | 39.6 (34.5, 45.2) |
| 25–29 | 44.9 (39.1, 51.4) | 42.2 (36.6, 48.4) | 42.8 (37.2, 48.9) | 41.0 (35.7, 46.9) | 41.3 (36.1, 47.1) | 44.1 (38.7, 50.0) |
| 30–39 | 60.8 (55.8, 66.2) | 61.3 (56.2, 66.6) | 57.2 (52.4, 62.3) | 61.5 (56.6, 66.7) | 59.2 (54.5, 64.2) | 54.2 (49.8, 58.9) |
| 40–49 | 83.8 (78.0, 89.9) | 81.8 (76.0, 87.9) | 80.7 (75.0, 86.8) | 80.1 (74.4, 86.2) | 76.7 (71.1, 82.6) | 78.9 (73.2, 84.9) |
| 50–59 | 87.7 (82.0, 93.6) | 97.4 (91.5, 104) | 94.8 (89.0, 101) | 94.2 (88.4, 100) | 96.7 (90.8, 103) | 99.4 (93.3, 106) |
| 60–69 | 114 (107, 122) | 112 (105, 120) | 106 (99.1, 114) | 107 (100, 115) | 102 (96.0, 109) | 107 (100, 114) |
| 70–79 | 126 (116, 138) | 116 (106, 127) | 123 (113, 133) | 120 (111, 131) | 123 (114, 133) | 114 (105, 123) |
| ≥80 | 121 (107, 137) | 115 (101, 129) | 119 (106, 134) | 127 (113, 141) | 126 (112, 140) | 113 (101, 127) |

Supplemental Table S3: Annual background rates of hospitalizations and emergency department visits for idiopathic thrombocytopenia by age group and sex in Ontario, 2015 to 2020

| Age group, years | Incidence rate per 100,000 population (95% confidence interval) | | | | | |
|---|---|---|---|---|---|---|
| | **2015** | **2016** | **2017** | **2018** | **2019** | **2020** |
| **Idiopathic thrombocytopenia, narrow definition** | | | | | | |
| **Both sexes** | | | | | | |
| 0–4 | 11.9 (9.51, 14.7) | 12.4 (9.95, 15.3) | 12.8 (10.3, 15.7) | 11.5 (9.19, 14.3) | 9.16 (7.08, 11.7) | 4.56 (3.14, 6.41) |
| 5–11 | 6.65 (5.18, 8.40) | 4.33 (3.17, 5.77) | 6.27 (4.86, 7.96) | 4.09 (2.97, 5.49) | 4.81 (3.60, 6.31) | 2.87 (1.95, 4.08) |
| 12–15 | 2.77 (1.61, 4.44) | 4.22 (2.76, 6.19) | 4.70 (3.15, 6.75) | 4.99 (3.39, 7.08) | 5.25 (3.61, 7.37) | 3.62 (2.29, 5.43) |
| 16–19 | 4.25 (2.85, 6.11) | 3.95 (2.61, 5.75) | 3.33 (2.11, 4.99) | 2.71 (1.63, 4.23) | 3.99 (2.65, 5.77) | 2.59 (1.54, 4.10) |
| 20–24 | 4.65 (3.38, 6.24) | 2.51 (1.61, 3.74) | 3.07 (2.07, 4.39) | 2.08 (1.29, 3.19) | 3.00 (2.04, 4.26) | 2.31 (1.48, 3.43) |
| 25–29 | 3.13 (2.10, 4.50) | 3.59 (2.49, 5.02) | 2.68 (1.75, 3.92) | 2.67 (1.76, 3.89) | 2.67 (1.78, 3.86) | 2.97 (2.03, 4.19) |
| **Females** | | | | | | |
| 0–4 | 10.9 (7.71, 15.0) | 7.69 (5.07, 11.2) | 11.7 (8.41, 15.9) | 8.56 (5.78, 12.2) | 7.70 (5.07, 11.2) | 5.11 (3.03, 8.08) |
| 5–11 | 5.61 (3.76, 8.06) | 4.79 (3.10, 7.07) | 4.95 (3.24, 7.26) | 4.16 (2.61, 6.30) | 4.34 (2.75, 6.51) | 3.22 (1.87, 5.15) |
| 12–15 | 3.68 (1.84, 6.59) | 5.64 (3.29, 9.04) | 4.30 (2.29, 7.35) | 4.60 (2.51, 7.72) | 4.87 (2.72, 8.02) | 4.49 (2.45, 7.53) |
| 16–19 | 4.24 (2.32, 7.11) | 5.14 (2.99, 8.23) | 4.48 (2.51, 7.39) | 3.24 (1.62, 5.79) | 5.28 (3.13, 8.35) | 3.55 (1.83, 6.20) |
| 20–24 | 6.39 (4.28, 9.18) | 2.41 (1.20, 4.31) | 3.21 (1.80, 5.30) | 2.70 (1.44, 4.62) | 4.07 (2.48, 6.28) | 3.63 (2.15, 5.73) |
| 25–29 | 4.60 (2.85, 7.03) | 4.97 (3.15, 7.46) | 3.38 (1.93, 5.48) | 3.26 (1.86, 5.29) | 3.94 (2.40, 6.08) | 3.84 (2.34, 5.92) |
| **Males** | | | | | | |
| 0–4 | 12.9 (9.46, 17.1) | 16.9 (13.0, 21.7) | 13.9 (10.3, 18.3) | 14.4 (10.8, 18.8) | 10.5 (7.50, 14.4) | 4.04 (2.26, 6.67) |
| 5–11 | 7.64 (5.48, 10.4) | 3.88 (2.40, 5.94) | 7.53 (5.41, 10.2) | 4.01 (2.51, 6.07) | 5.27 (3.53, 7.57) | 2.55 (1.39, 4.27) |
| 12–15 | 1.91 (0.70, 4.15) | 2.86 (1.31, 5.43) | 5.08 (2.90, 8.25) | 5.36 (3.12, 8.58) | 5.62 (3.33, 8.88) | 2.78 (1.27, 5.28) |
| 16–19 | 4.26 (2.39, 7.03) | 2.84 (1.36, 5.22) | 2.24 (0.97, 4.41) | 2.21 (0.95, 4.36) | 2.78 (1.33, 5.11) | 1.69 (0.62, 3.67) |
| 20–24 | 3.05 (1.71, 5.03) | 2.61 (1.39, 4.47) | 2.95 (1.65, 4.86) | 1.52 (0.66, 2.99) | 2.03 (1.02, 3.64) | 1.10 (0.41, 2.40) |
| 25–29 | 1.70 (0.74, 3.36) | 2.28 (1.14, 4.07) | 2.01 (0.96, 3.69) | 2.12 (1.06, 3.79) | 1.48 (0.64, 2.92) | 2.16 (1.12, 3.77) |
| **Idiopathic thrombocytopenia, broad definition** | | | | | | |
| **Both sexes** | | | | | | |
| All ages | 40.5 (39.4, 41.5) | 43.5 (42.4, 44.6) | 44.8 (43.7, 45.9) | 44.9 (43.8, 46.0) | 45.8 (44.8, 47.0) | 46.5 (45.4, 47.6) |
| 0–4 | 31.7 (27.7, 36.1) | 29.1 (25.3, 33.3) | 34.5 (30.3, 39.0) | 31.1 (27.0, 35.5) | 30.0 (26.1, 34.3) | 26.3 (22.7, 30.3) |
| 5–11 | 16.2 (13.9, 18.9) | 12.3 (10.3, 14.6) | 13.8 (11.7, 16.3) | 12.4 (10.4, 14.7) | 13.1 (11.0, 15.4) | 10.2 (8.38, 12.3) |
| 12–15 | 11.4 (8.90, 14.4) | 10.9 (8.43, 13.8) | 14.4 (11.6, 17.7) | 14.2 (11.4, 17.5) | 13.0 (10.4, 16.2) | 12.1 (9.56, 15.1) |
| 16–19 | 13.8 (11.1, 16.9) | 12.6 (10.1, 15.6) | 13.3 (10.7, 16.3) | 10.7 (8.41, 13.4) | 14.8 (12.0, 17.8) | 11.1 (8.76, 13.9) |
| 20–24 | 13.0 (10.8, 15.5) | 9.43 (7.58, 11.6) | 11.5 (9.45, 13.8) | 9.53 (7.72, 11.6) | 10.1 (8.23, 12.2) | 8.85 (7.13, 10.9) |
| 25–29 | 13.2 (10.9, 15.7) | 14.5 (12.2, 17.1) | 13.0 (10.8, 15.4) | 13.0 (10.8, 15.4) | 13.9 (11.8, 16.4) | 14.9 (12.7, 17.4) |
| 30–39 | 15.1 (13.4, 17.0) | 18.1 (16.2, 20.1) | 18.0 (16.1, 20.0) | 19.6 (17.7, 21.7) | 19.1 (17.2, 21.1) | 19.7 (17.8, 21.7) |

| | | | | | | |
|---|---|---|---|---|---|---|
| 40–49 | 23.0 (20.9, 25.3) | 24.2 (22.0, 26.5) | 24.9 (22.7, 27.3) | 25.6 (23.4, 28.0) | 26.4 (24.1, 28.8) | 24.0 (21.8, 26.4) |
| 50–59 | 40.6 (37.9, 43.4) | 45.3 (42.5, 48.3) | 43.8 (41.0, 46.7) | 42.8 (40.1, 45.7) | 45.6 (42.7, 48.6) | 47.8 (44.9, 50.9) |
| 60–69 | 64.6 (60.6, 68.7) | 73.0 (68.8, 77.3) | 73.7 (69.6, 78.0) | 73.3 (69.2, 77.5) | 73.5 (69.5, 77.7) | 79.1 (75.0, 83.3) |
| 70–79 | 118 (111, 125) | 127 (120, 135) | 125 (118, 132) | 128 (121, 135) | 135.0 (128, 142) | 129 (123, 136) |
| ≥80 | 198 (186, 210) | 208 (197, 220) | 221 (210, 233) | 221 (210, 233) | 209 (198, 220) | 221 (210, 233) |
| **Females** | | | | | | |
| All ages | 34.9 (33.5, 36.3) | 37.7 (36.3, 39.2) | 39.4 (37.9, 40.9) | 38.5 (37.1, 40.0) | 39.8 (38.3, 41.2) | 39.3 (37.9, 40.8) |
| 0–4 | 26.1 (21.0, 32.0) | 19.9 (15.6, 25.2) | 32.0 (26.4, 38.5) | 25.1 (20.1, 30.9) | 26.5 (21.4, 32.5) | 27.8 (22.6, 33.9) |
| 5–11 | 13.7 (10.3, 17.3) | 11.3 (8.61, 14.6) | 10.1 (7.56, 13.2) | 12.3 (9.49, 15.7) | 11.7 (8.97, 15.0) | 10.2 (7.68, 13.3) |
| 12–15 | 11.4 (7.88, 15.9 | 12.0 (8.37, 16.6) | 13.6 (9.73, 18.4) | 11.2 (7.74, 15.6) | 14.3 (10.4, 19.2) | 14.4 (10.5, 19.3) |
| 16–19 | 15.7 (11.8, 20.6) | 15.4 (11.5, 20.3) | 13.2 (9.56, 17.7) | 11.2 (7.91, 15.4) | 17.0 (12.9, 22.0) | 11.8 (8.45, 16.1) |
| 20–24 | 15.0 (11.6, 19.0) | 11.6 (8.69, 15.2) | 12.8 (9.80, 16.5) | 10.6 (7.90, 14.0) | 11.4 (8.60, 14.8) | 9.87 (7.30, 13.1) |
| 25–29 | 16.2 (12.7, 20.4) | 17.7 (14.1 22.0) | 15.4 (12.1, 19.4) | 12.6 (9.67, 16.2) | 19.5 (15.8, 23.7) | 17.6 (14.2, 21.6) |
| 30–39 | 17.4 (14.1, 20.1) | 19.9 (17.1, 23.0) | 20.7 (17.9, 23.8) | 22.0 (19.2, 25.2) | 21.4 (18.6, 24.5) | 20.6 (17.9, 23.6) |
| 40–49 | 21.7 (18.9, 24.9) | 22.3 (19.4, 25.5) | 24.6 (21.6, 28.0) | 22.1 (19.2, 25.3) | 23.6 (20.6, 26.9) | 19.9 (17.1, 22.9) |
| 50–59 | 33.5 (30.1, 37.2) | 38.7 (35.0, 42.6) | 37.8 (34.2, 41.7) | 35.2 (31.7, 39.0) | 37.2 (33.6, 41.1) | 38.8 (35.1, 42.8) |
| 60–69 | 50.9 (46.0, 56.0) | 57.4 (52.3, 62.8) | 57.3 (52.3, 62.6) | 60.1 (55.0, 65.5) | 59.1 (54.2, 64.4) | 62.7 (57.7, 68.1) |
| 70–79 | 84.1 (76.1, 92.7) | 102.6 (93.9, 112) | 94.7 (86.6, 103) | 92.6 (84.7, 101) | 102 (94.0, 111) | 95.1 (87.5, 103) |
| ≥80 | 144 (132, 157) | 144 (132, 157) | 167 (154, 181) | 168 (155, 182) | 150 (138, 162) | 155 (143, 168) |
| **Males** | | | | | | |
| All ages | 46.2 (44.6, 47.9) | 49.4 (47.7, 51.1) | 50.4 (48.8, 52.1) | 51.3 (49.7, 53.0) | 52.1 (50.4, 53.8) | 53.8 (52.1, 55.5) |
| 0–4 | 37.0 (31.0, 43.8) | 37.9 (31.8, 44.7) | 36.8 (30.8, 43.5) | 36.9 (30.9, 43.6) | 33.3 (27.6, 39.7) | 24.8 (20.0, 30.4) |
| 5–11 | 18.6 (15.2, 22.7) | 13.3 (10.4, 16.8) | 17.5 (14.1, 21.3) | 12.6 (9.79, 15.9) | 14.4 (11.4, 17.9) | 10.2 (7.69, 13.2) |
| 12–15 | 11.5 (8.02, 15.9) | 9.85 (6.70, 14.0) | 15.2 (11.2, 20.2) | 17.0 (12.8, 22.2) | 11.9 (8.39, 16.3) | 9.88 (6.76, 14.0) |
| 16–19 | 11.9 (8.60, 16.1) | 9.94 (6.92, 13.8) | 13.4 (9.91, 17.8) | 10.2 (7.20, 14.1) | 12.5 (9.11, 16.7) | 10.4 (7.33, 14.3) |
| 20–24 | 11.2 (8.42, 14.5) | 7.44 (5.24, 10.3) | 10.2 (7.63, 13.4) | 8.54 (6.23, 11.4) | 8.87 (6.54, 11.8) | 7.92 (5.73, 10.7) |
| 25–29 | 10.2 (7.54, 13.6) | 11.4 (8.57, 14.8) | 10.7 (7.97, 13.9) | 13.3 (10.3, 16.8) | 8.71 (6.40, 11.6) | 12.4 (9.66, 15.7) |
| 30–39 | 12.7 (10.4, 15.3) | 16.2 (13.7, 19.0) | 15.2 (12.7, 17.9) | 17.1 (14.6, 20.0) | 16.8 (14.3, 19.5) | 18.7 (16.2, 21.6) |
| 40–49 | 24.4 (21.3, 27.8) | 26.2 (23.0, 29.7) | 25.2 (22.0, 28.7) | 29.3 (25.9, 33.0) | 29.3 (25.9, 33.1) | 28.4 (25.0, 32.1) |
| 50–59 | 47.7 (43.6, 52.1) | 52.1 (47.8, 56.7) | 49.8 (45.6, 54.3) | 50.6 (46.3, 55.1) | 54.1 (49.7, 58.8) | 57.1 (52.5, 61.9) |
| 60–69 | 79.3 (73.0, 86.0) | 89.8 (83.2, 96.8) | 91.4 (84.8, 98.3) | 87.5 (81.1, 94.2) | 88.9 (82.6, 95.5) | 96.5 (90.1, 103) |
| 70–79 | 157 (145, 169) | 156 (144, 168) | 159.6 (148.3, 172) | 168 (156, 180) | 173 (161, 185) | 169 (158, 180) |
| ≥80 | 282 (261, 305) | 308 (286, 332) | 304 (282, 327) | 302 (281, 324) | 298 (277, 320) | 320 (299, 342) |

Supplemental Table S4: Annual background rates of hospitalizations and emergency department visits for febrile convulsions by age group and sex in Ontario, 2015 to 2020

| Age group, years | Incidence rate per 100,000 population (95% confidence interval) | | | | | |
|---|---|---|---|---|---|---|
| | **2015** | **2016** | **2017** | **2018** | **2019** | **2020** |
| **Both sexes** | | | | | | |
| All ages | 23.5 (22.7, 24.3) | 28.6 (27.7, 29.5) | 23.7 (22.9, 24.5) | 25.5 (24.6, 26.3) | 24.0 (23.2, 24.8) | 13.5 (12.9, 14.1) |
| 0–4 | 418 (403, 434) | 504 (487, 520) | 428 (413, 444) | 468 (452, 484) | 449 (433, 464) | 250 (239, 262) |
| 5–11 | 17.5 (15.0, 20.2) | 25.0 (22.1, 28.2) | 17.9 (15.4, 20.6) | 21.0 (18.3, 23.9) | 19.2 (16.6, 22.0) | 11.7 (9.73, 13.9) |
| 12–15 | 1.63 (0.78, 3.00) | 2.92 (1.73, 4.62) | 0.49 (0.10, 1.42) | 0.80 (0.26, 1.88) | 0.95 (0.35, 2.08) | 0.31 (0.04, 1.14) |
| 16–19 | 0.73 (0.24, 1.71) | 1.03 (0.41, 2.11) | 0.58 (0.16, 1.48) | 0.57 (0.16, 1.46) | 0.43 (0.09, 1.25) | 0.43 (0.09, 1.26) |
| 20–24 | 0.21 (0.03, 0.76) | 0.52 (0.17, 1.22) | 0.72 (0.29, 1.48) | 0.20 (0.02, 0.72) | 0.29 (0.06, 0.85) | 0.19 (0.02, 0.69) |
| 25–29 | 0.11 (0.00, 0.60) | 0.42 (0.12, 1.08) | 0.51 (0.17, 1.20) | 0.49 (0.16, 1.15) | 0.19 (0.02, 0.69) | 0.09 (0.00, 0.52) |
| 30–39 | 0.22 (0.06, 0.57) | 0.49 (0.23, 0.94) | 0.27 (0.09, 0.63) | 0.26 (0.09, 0.61) | 0.20 (0.06, 0.52) | 0.34 (0.14, 0.71) |
| 40–49 | 0.16 (0.03, 0.47) | 0.27 (0.09, 0.62) | 0.48 (0.22, 0.92) | 0.49 (0.22, 0.92) | 0.27 (0.09, 0.63) | 0.27 (0.09, 0.63) |
| 50–59 | 0.39 (0.17, 0.76) | 0.43 (0.20, 0.82) | 0.53 (0.26, 0.95) | 0.24 (0.08, 0.56) | 0.39 (0.17, 0.77) | 0.44 (0.20, 0.84) |
| 60–69 | 0.32 (0.10, 0.75) | 0.69 (0.34, 1.23) | 0.55 (0.25, 1.05) | 0.24 (0.07, 0.61) | 0.58 (0.28, 1.07) | 0.68 (0.35, 1.19) |
| 70–79 | 0.34 (0.07, 0.98) | 0.75 (0.30, 1.55) | 1.01 (0.49, 1.86) | 0.48 (0.16, 1.12) | 0.46 (0.15, 1.07) | 0.26 (0.05, 0.77) |
| ≥80 | 1.21 (0.49, 2.50) | 0.84 (0.27, 1.97) | 1.64 (0.79, 3.02) | 0.96 (0.35, 2.09) | 1.25 (0.54, 2.46) | 0.91 (0.34, 1.99) |
| **Females** | | | | | | |
| All ages | 20.2 (19.2, 21.3) | 24.3 (23.2, 25.5) | 20.7 (19.6, 21.8) | 21.9 (20.9, 23.0) | 20.1 (19.1, 21.2) | 12.5 (11.7, 13.3) |
| 0–4 | 377 (357, 398) | 450 (428, 473) | 391 (371, 412) | 420 (399, 442) | 393 (372, 414) | 239 (223, 255) |
| 5–11 | 13.2 (10.2, 16.7) | 18.2 (14.7, 22.3) | 13.3 (10.4, 16.9) | 18.0 (14.6, 22.0) | 14.0 (11.0, 17.5) | 11.5 (8.83, 14.8) |
| 12–15 | 1.34 (0.36, 3.43) | 2.99 (1.37, 5.67) | 0.00 (0.00, 0.00) | 0.66 (0.08, 2.37) | 0.32 (0.01, 1.81) | 0.00 (0.00, 0.00) |
| 16–19 | 0.30 (0.01, 1.69) | 0.60 (0.07, 2.18) | 0.30 (0.01, 1.67) | 0.59 (0.07, 2.13) | 0.29 (0.01, 1.64) | 0.89 (0.18, 2.59) |
| 20–24 | 0.22 (0.01, 1.23) | 0.44 (0.05, 1.58) | 1.28 (0.47, 2.80) | 0.00 (0.00, 0.00) | 0.00 (0.00, 0.00) | 0.00 (0.00, 0.00) |
| 25–29 | 0.22 (0.01, 1.22) | 0.65 (0.13, 1.90) | 0.63 (0.13, 1.85) | 0.41 (0.05, 1.47) | 0.20 (0.00, 1.10) | 0.19 (0.00, 1.07) |
| 30–39 | 0.43 (0.12, 1.11) | 0.54 (0.17, 1.25) | 0.11 (0.00, 0.59) | 0.21 (0.03, 0.75) | 0.20 (0.02, 0.73) | 0.59 (0.22, 1.29) |
| 40–49 | 0.10 (0.00, 0.58) | 0.31 (0.06, 0.92) | 0.42 (0.11, 1.08) | 0.53 (0.17, 1.23) | 0.32 (0.07, 0.92) | 0.32 (0.06, 0.92) |
| 50–59 | 0.29 (0.06, 0.84) | 0.38 (0.10, 0.98) | 0.57 (0.21, 1.25) | 0.10 (0.00, 0.53) | 0.48 (0.16, 1.12) | 0.49 (0.16, 1.13) |
| 60–69 | 0.37 (0.08, 1.09) | 0.24 (0.03, 0.87) | 0.35 (0.07, 1.04) | 0.12 (0.00, 0.64) | 0.56 (0.18, 1.31) | 0.44 (0.12, 1.12) |
| 70–79 | 0.21 (0.01, 1.16) | 0.81 (0.22, 2.07) | 0.95 (0.31, 2.21) | 0.54 (0.11, 1.58) | 0.34 (0.04, 1.24) | 0.17 (0.00, 0.92) |
| ≥80 | 1.42 (0.46, 3.30) | 0.55 (0.07, 2.00) | 1.90 (0.76, 3.92) | 0.53 (0.06, 1.92) | 1.82 (0.73, 3.75) | 1.02 (0.28, 2.61) |
| **Males** | | | | | | |

| | | | | | | |
|---|---|---|---|---|---|---|
| All ages | 26.8 (25.6, 28.1) | 32.9 (31.6, 34.3) | 26.8 (25.6, 28.1) | 29.1 (27.8, 30.4) | 28.1 (26.8, 29.3 | 14.5 (13.6, 15.4) |
| 0–4 | 457 (436, 480) | 555 (531, 579) | 463 (442, 486) | 513 (490, 537) | 502 (479, 525) | 261 (244, 278) |
| 5–11 | 21.6 (17.9, 25.9) | 31.6 (27.1, 36.7) | 22.2 (18.5, 26.6) | 23.9 (20.0, 28.4) | 24.2 (20.2, 28.6) | 11.8 (9.12, 15.1) |
| 12–15 | 1.91 (0.70, 4.15) | 2.86 (1.31, 5.43) | 0.95 (0.20, 2.78) | 0.95 (0.20, 2.77) | 1.56 (0.51, 3.64) | 0.62 (0.07, 2.23) |
| 16–19 | 1.14 (0.31, 2.91) | 1.42 (0.46, 3.31) | 0.84 (0.17, 2.46) | 0.55 (0.07, 2.00) | 0.56 (0.07, 2.01) | 0.00 (0.00, 0.00) |
| 20–24 | 0.20 (0.01, 1.13) | 0.60 (0.12, 1.76) | 0.20 (0.00, 1.09) | 0.38 (0.05, 1.37) | 0.55 (0.11, 1.62) | 0.37 (0.04, 1.33) |
| 25–29 | 0.00 (0.00, 0.00) | 0.21 (0.01, 1.15) | 0.40 (0.05, 1.45) | 0.58 (0.12, 1.69) | 0.19 (0.00, 1.03) | 0.00 (0.00, 0.00) |
| 30–39 | 0.00 (0.00, 0.00) | 0.45 (0.12, 1.14) | 0.44 (0.12, 1.12) | 0.32 (0.07, 0.93) | 0.20 (0.02, 0.73) | 0.10 (0.00, 0.55) |
| 40–49 | 0.22 (0.03, 0.78) | 0.22 (0.03, 0.79) | 0.55 (0.18, 1.28) | 0.44 (0.12, 1.13) | 0.22 (0.03, 0.80) | 0.22 (0.03, 0.80) |
| 50–59 | 0.49 (0.16, 1.13) | 0.48 (0.16, 1.13) | 0.48 (0.16, 1.13) | 0.39 (0.11, 1.00) | 0.29 (0.06, 0.86) | 0.40 (0.11, 1.01) |
| 60–69 | 0.27 (0.03, 0.97) | 1.17 (0.54, 2.22) | 0.76 (0.28, 1.66) | 0.37 (0.08, 1.09) | 0.60 (0.20, 1.41) | 0.94 (0.40, 1.85) |
| 70–79 | 0.48 (0.06, 1.74) | 0.69 (0.14, 2.03) | 1.09 (0.35, 2.54) | 0.41 (0.05, 1.49) | 0.59 (0.12, 1.73) | 0.38 (0.05, 1.37) |
| ≥80 | 0.89 (0.11, 3.22) | 1.29 (0.27, 3.77) | 1.25 (0.26, 3.64) | 1.61 (0.44, 4.12) | 0.39 (0.01, 2.17) | 0.76 (0.09, 2.74) |

Supplemental Table S5: Annual background rates of hospitalizations and emergency department visits for acute disseminated encephalomyelitis by age group and sex in Ontario, 2015 to 2020

| Age group, years | Incidence rate per 100,000 population (95% confidence interval) | | | | | |
|---|---|---|---|---|---|---|
| | 2015 | 2016 | 2017 | 2018 | 2019 | 2020 |
| **Acute disseminated encephalomyelitis, narrow definition*** | | | | | | |
| **Both sexes** | | | | | | |
| All ages | 0.16 (0.10, 0.24) | 0.22 (0.15, 0.31) | 0.22 (0.15, 0.31) | 0.20 (0.13, 0.28) | 0.14 (0.09, 0.22) | 0.10 (0.06, 0.17) |
| 0–4 | 0.56 (0.15, 1.44) | 0.42 (0.09, 1.22) | 0.28 (0.03, 1.01) | 0.14 (0.00, 0.77) | 1.11 (0.48, 2.19) | 0.41 (0.09, 1.21) |
| 5–11 | 0.66 (0.27, 1.37) | 0.47 (0.15, 1.10) | 0.75 (0.32, 1.47) | 0.09 (0.00, 0.52) | 0.19 (0.02, 0.67) | 0.00 (0.00, 0.00) |
| 12–15 | 0.16 (0.00, 0.91) | 0.32 (0.04, 1.17) | 0.32 (0.04, 1.17) | 0.32 (0.04, 1.16) | 0.16 (0.00, 0.89) | 0.00 (0.00, 0.00) |
| 16–19 | 0.29 (0.04, 1.06) | 0.29 (0.04, 1.06) | 0.29 (0.04, 1.04) | 0.29 (0.03, 1.03) | 0.00 (0.00, 0.00) | 0.00 (0.00, 0.00) |
| 20–24 | 0.00 (0.00, 0.00) | 0.00 (0.00, 0.00) | 0.31 (0.06, 0.90) | 0.20 (0.02, 0.72) | 0.19 (0.02, 0.70) | 0.00 (0.00, 0.00) |
| 25–29 | 0.00 (0.00, 0.00) | 0.53 (0.17, 1.23) | 0.10 (0.00, 0.57) | 0.00 (0.00, 0.00) | 0.10 (0.00, 0.53) | 0.09 (0.00, 0.52) |
| 30–39 | 0.06 (0.00, 0.31) | 0.11 (0.01, 0.40) | 0.11 (0.01, 0.39) | 0.21 (0.06, 0.54) | 0.00 (0.00, 0.00) | 0.15 (0.03, 0.43) |
| 40–49 | 0.11 (0.01, 0.38) | 0.11 (0.01, 0.39) | 0.05 (0.00, 0.30) | 0.16 (0.03, 0.47) | 0.05 (0.00, 0.30) | 0.05 (0.00, 0.30) |
| 50–59 | 0.14 (0.03, 0.42) | 0.29 (0.11, 0.63) | 0.19 (0.05, 0.49) | 0.05 (0.00, 0.27) | 0.15 (0.03, 0.43) | 0.15 (0.03, 0.43) |
| 60–69 | 0.06 (0.00, 0.36) | 0.06 (0.00, 0.35) | 0.37 (0.13, 0.80) | 0.48 (0.21, 0.94) | 0.00 (0.00, 0.00) | 0.11 (0.01, 0.41) |
| 70–79 | 0.11 (0.00, 0.62) | 0.22 (0.03, 0.78) | 0.00 (0.00, 0.00) | 0.10 (0.00, 0.54) | 0.28 (0.06, 0.81) | 0.18 (0.02, 0.64) |
| ≥80 | 0.00 (0.00, 0.00) | 0.00 (0.00, 0.00) | 0.00 (0.00, 0.00) | 0.48 (0.10, 1.40) | 0.00 (0.00, 0.00) | 0.00 (0.00, 0.00) |
| **Acute disseminated encephalomyelitis, broad definition** | | | | | | |
| **Both sexes** | | | | | | |
| All ages | 21.8 (21.1, 22.6) | 23.0 (22.2, 23.8) | 23.2 (22.4, 24.0) | 23.0 (22.2, 23.8) | 22.9 (22.1, 23.7) | 20.1 (19.4, 20.8) |
| 0–4 | 8.97 (6.91, 11.5) | 7.38 (5.53, 9.65) | 7.53 (5.66, 9.83) | 9.0 (6.97, 11.5) | 11.0 (8.68, 13.7) | 8.99 (6.94, 11.5) |
| 5–11 | 4.46 (3.28, 5.93) | 3.01 (2.06, 4.25) | 4.30 (3.15, 5.74) | 3.90 (2.81, 5.27) | 4.63 (3.44, 6.10) | 3.52 (2.49, 4.84) |
| 12–15 | 4.57 (3.03, 6.60) | 4.87 (3.29, 6.96) | 4.21 (2.75, 6.17) | 4.99 (3.39, 7.08) | 4.93 (3.35, 7.00) | 5.98 (4.23, 8.20) |
| 16–19 | 5.28 (3.70, 7.31) | 7.18 (5.31, 9.49) | 6.36 (4.62, 8.54) | 5.84 (4.19, 7.93) | 8.27 (6.28, 10.7) | 5.33 (3.76, 7.35) |
| 20–24 | 7.19 (5.58, 9.11) | 8.07 (6.37, 10.1) | 9.32 (7.51, 11.5) | 8.54 (6.83, 10.5) | 7.46 (5.88, 9.32) | 7.02 (5.50, 8.83) |
| 25–29 | 13.1 (10.8, 15.6) | 13.7 (11.5, 16.3) | 14.1 (11.8, 16.7) | 13.4 (11.2, 15.8) | 13.1 (11.0, 15.5) | 9.93 (8.14, 12.0) |
| 30–39 | 19.6 (17.6, 21.8) | 19.7 (17.7, 21.8) | 19.0 (17.1, 21.1) | 19.3 (17.4, 21.4) | 20.8 (18.8, 22.9) | 17.3 (15.5, 19.2) |
| 40–49 | 23.1 (21.0, 25.4) | 25.4 (23.2, 27.8) | 25.3 (23.1, 27.7) | 27.2 (24.9, 29.7) | 25.0 (22.8, 27.4) | 22.9 (20.8, 25.2) |
| 50–59 | 32.7 (30.3, 35.2) | 35.9 (33.3, 38.5) | 32.1 (29.7, 34.6) | 31.9 (29.5, 34.4) | 33.8 (31.4, 36.4) | 27.7 (25.5, 30.1) |
| 60–69 | 38.5 (35.5, 41.7) | 39.7 (36.7, 42.9) | 41.7 (38.7, 45.0) | 39.9 (36.9, 43.1) | 38.2 (35.3, 41.2) | 34.7 (32.0, 37.5) |
| 70–79 | 41.9 (37.8, 46.4) | 43.8 (39.6, 48.2) | 46.2 (42.1, 50.6) | 44.8 (40.8, 49.1) | 41.1 (37.4, 45.1) | 38.9 (35.3, 42.7) |
| ≥80 | 33.3 (28.7, 38.3) | 34.0 (29.5, 39.1) | 39.4 (34.6, 44.7) | 36.2 (31.6, 41.2) | 35.1 (30.7, 40.0) | 31.7 (27.6, 36.3) |
| **Females** | | | | | | |

| | | | | | | |
|---|---|---|---|---|---|---|
| All ages | 25.9 (24.7, 27.1) | 27.3 (26.1, 28.6) | 27.0 (25.8, 28.2) | 26.4 (25.3, 27.7) | 26.6 (25.5, 27.9) | 22.7 (21.7, 23.8) |
| 0–4 | 9.75 (6.75, 13.6) | 7.98 (5.30, 11.5) | 7.43 (4.85, 10.9) | 7.99 (5.31, 11.6) | 9.69 (6.71, 13.5) | 7.10 (4.60, 10.5) |
| 5–11 | 4.06 (2.52, 6.21) | 3.07 (1.75, 4.98) | 3.81 (2.33, 5.88) | 4.73 (3.06, 6.98) | 4.53 (2.90, 6.74) | 3.03 (1.73, 4.92) |
| 12–15 | 5.35 (3.06, 8.69) | 5.64 (3.29, 9.04) | 3.97 (2.05, 6.93) | 5.26 (3.00, 8.54) | 4.87 (2.72, 8.02) | 6.41 (3.92, 9.90) |
| 16–19 | 6.36 (3.93, 9.72) | 10.3 (7.12, 14.4) | 7.77 (5.08, 11.4) | 6.18 (3.83, 9.45) | 11.4 (8.14, 15.7) | 6.80 (4.31, 10.2) |
| 20–24 | 10.4 (7.61, 13.8) | 10.3 (7.56, 13.7) | 11.1 (8.31, 14.6) | 12.5 (9.53, 16.1) | 10.4 (7.72, 13.6) | 8.86 (6.44, 11.9) |
| 25–29 | 16.2 (12.7, 20.4) | 20.8 (16.8, 25.3) | 20.1 (16.2, 24.5) | 18.1 (14.5, 22.3) | 16.9 (13.5, 20.9) | 13.2 (10.3, 16.8) |
| 30–39 | 26.0 (22.8, 29.5) | 25.9 (22.7, 29.4) | 24.5 (21.5, 27.9) | 24.3 (21.3, 27.7) | 25.8 (22.8, 29.2) | 20.7 (18.0, 23.7) |
| 40–49 | 28.8 (25.5, 32.4) | 31.9 (28.4, 35.7) | 31.2 (27.7, 34.9) | 34.5 (30.8, 38.4) | 32.5 (29.0, 36.3) | 29.6 (26.3, 33.3) |
| 50–59 | 38.9 (35.2, 42.9) | 43.3 (39.4, 47.5) | 38.4 (34.7, 42.4) | 35.0 (31.5, 38.8) | 40.1 (36.3, 44.1) | 31.0 (27.7, 34.5) |
| 60–69 | 42.4 (38.0, 47.1) | 44.7 (40.3, 49.5) | 45.4 (41.0, 50.2) | 43.7 (39.4, 48.3) | 41.1 (37.0, 45.5) | 37.3 (33.4, 41.5) |
| 70–79 | 47.6 (41.6, 54.2) | 42.5 (37.0, 48.7) | 46.5 (40.9, 52.7) | 45.6 (40.1, 51.5) | 43.2 (38.0, 48.9) | 37.8 (33.1, 43.0) |
| ≥80 | 28.3 (23.0, 34.4) | 29.1 (23.8, 35.2) | 36.7 (30.3, 43.4) | 31.9 (26.5, 38.2) | 30.2 (24.9, 36.2) | 29.8 (24.7, 35.8) |
| **Males** | | | | | | |
| All ages | 17.6 (16.7, 18.7) | 18.6 (17.6, 19.6) | 19.4 (18.4, 20.4) | 19.5 (18.5, 20.6) | 19.1 (18.1, 20.1) | 17.4 (16.5, 18.4) |
| 0–4 | 8.22 (5.55, 11.7) | 6.81 (4.41, 10.1) | 7.63 (5.07, 11.0) | 10.0 (7.06, 13.8) | 12.2 (8.9, 16.3) | 10.8 (7.70, 14.7) |
| 5–11 | 4.85 (3.17, 7.10) | 2.96 (1.69, 4.81) | 4.78 (3.12, 7.00) | 3.10 (1.81, 4.96) | 4.72 (3.09, 6.92) | 4.00 (2.51, 6.06) |
| 12–15 | 3.82 (1.97, 6.67) | 4.13 (2.20, 7.07) | 4.44 (2.43, 7.45) | 4.73 (2.65, 7.80) | 4.99 (2.85, 8.11) | 5.56 (3.29, 8.79) |
| 16–19 | 4.26 (2.39, 7.03) | 4.26 (2.38, 7.03) | 5.04 (2.99, 7.97) | 5.53 (3.38, 8.54) | 5.28 (3.18, 8.24) | 3.94 (2.15, 6.61) |
| 20–24 | 4.27 (2.64, 6.52) | 6.03 (4.07, 8.61) | 7.66 (5.45, 10.5) | 4.94 (3.22, 7.23) | 4.81 (3.14, 7.04) | 5.34 (3.58, 7.67) |
| 25–29 | 10.0 (7.35, 13.3) | 7.03 (4.87, 9.83) | 8.44 (6.08, 11.4) | 8.85 (6.48, 11.8) | 9.45 (7.04, 12.4) | 6.84 (4.84, 9.38) |
| 30–39 | 12.9 (10.6, 15.5) | 13.2 (10.9, 15.8) | 13.3 (11.1, 15.9) | 14.3 (12.0, 16.9) | 15.8 (13.4, 18.4) | 13.8 (11.6, 16.3) |
| 40–49 | 17.3 (14.7, 20.1) | 18.7 (16.0, 21.7) | 19.3 (16.5, 22.3) | 19.7 (16.9, 22.8) | 17.2 (14.6, 20.1) | 15.8 (13.3, 18.6) |
| 50–59 | 26.3 (23.3, 29.7) | 28.3 (25.2, 31.7) | 25.7 (22.7, 29.0) | 28.7 (25.5, 32.2) | 27.5 (24.4, 30.9) | 24.4 (21.5, 27.7) |
| 60–69 | 34.4 (30.3, 38.8) | 34.4 (30.3, 38.8) | 37.8 (33.6, 42.4) | 35.9 (31.8, 40.2) | 35.1 (31.2, 39.4) | 31.9 (28.2, 35.9) |
| 70–79 | 35.4 (29.9, 41.6) | 45.2 (39.0, 52.0) | 45.9 (39.9, 52.5) | 43.9 (38.2, 50.2) | 38.8 (33.6, 44.6) | 40.1 (34.9, 45.9) |
| ≥80 | 41.0 (33.1, 50.3) | 41.7 (33.8, 50.8) | 43.6 (35.7, 52.8) | 42.6 (34.9, 51.6) | 42.4 (34.9, 51.2) | 34.5 (27.8, 42.4) |

* Separate rates for females and males could not be generated for the narrow definition because of small counts.

Supplemental Table S6: Annual background rates of hospitalizations and emergency department visits for myocarditis and pericarditis by age group and sex in Ontario, 2015 to 2020

| Age group, years | Incidence rate per 100,000 population (95% confidence interval) | | | | | |
|---|---|---|---|---|---|---|
| | **2015** | **2016** | **2017** | **2018** | **2019** | **2020** |
| **Myocarditis/pericarditis** | | | | | | |
| **Both sexes** | | | | | | |
| All ages | 9.82 (9.30, 10.4) | 10.2 (9.69, 10.8) | 12.4 (11.8, 13.0) | 11.9 (11.3, 12.4) | 12.0 (11.4, 12.6) | 10.7 (10.2, 11.2) |
| 0–4 | 0.56 (0.15, 1.44) | 1.67 (0.86, 2.92) | 1.12 (0.48, 2.20) | 1.53 (0.76, 2.74) | 1.25 (0.57, 2.37) | 0.55 (0.15, 1.42) |
| 5–11 | 0.76 (0.33, 1.50) | 1.22 (0.65, 2.09) | 0.94 (0.45, 1.72) | 0.74 (0.32, 1.46) | 0.46 (0.15, 1.08) | 0.74 (0.32, 1.46) |
| 12–15 | 0.98 (0.36, 2.13) | 2.76 (1.61, 4.42) | 3.40 (2.11, 5.20) | 2.74 (1.59, 4.38) | 2.07 (1.10, 3.54) | 2.36 (1.32, 3.89) |
| 16–19 | 9.68 (7.48, 12.3) | 9.08 (6.96, 11.6) | 16.1 (13.2, 19.3) | 11.4 (9.04, 14.2) | 13.4 (10.8, 16.4) | 7.50 (5.60, 9.83) |
| 20–24 | 12.6 (10.4, 15.1) | 11.8 (9.76, 14.2) | 13.2 (11.0, 15.7) | 12.4 (10.3, 14.8) | 12.9 (10.8, 15.6) | 8.95 (7.22, 11.0) |
| 25–29 | 8.85 (7.04, 11.0) | 11.0 (8.98, 13.3) | 13.0 (10.8, 15.4) | 11.7 (9.66, 14.0) | 11.3 (9.32, 13.5) | 9.28 (7.55, 11.3) |
| 30–39 | 11.1 (9.60, 12.7) | 10.9 (9.43, 12.5) | 11.7 (10.2, 13.3) | 12.2 (10.6, 13.8) | 12.0 (10.5, 13.6) | 11.0 (9.61, 12.6) |
| 40–49 | 9.92 (8.55, 11.5) | 10.8 (9.37, 12.4) | 12.8 (11.2, 14.5) | 12.8 (11.3, 14.6) | 11.8 (10.3, 13.4) | 11.1 (9.60, 12.7) |
| 50–59 | 11.5 (10.1, 13.1) | 12.2 (10.7, 13.8) | 13.8 (12.3, 15.5) | 13.4 (11.9, 15.1) | 12.9 (11.4, 14.6) | 12.7 (11.2, 14.4) |
| 60–69 | 13.1 (11.4, 15.1) | 13.3 (11.6, 15.2) | 16.5 (14.6, 18.6) | 16.2 (14.3, 18.3) | 17.0 (15.1, 19.0) | 15.6 (13.8, 17.5) |
| 70–79 | 15.2 (12.8, 18.0) | 16.1 (13.6, 18.9) | 21.6 (18.8, 24.7) | 19.9 (17.3, 22.8) | 21.4 (18.7, 24.3) | 18.3 (15.9, 21.0) |
| ≥80 | 16.8 (13.6, 20.5) | 13.6 (10.8, 17.0) | 18.7 (15.4, 22.5) | 17.9 (14.8, 21.6) | 19.7 (16.4, 23.4) | 20.0 (16.7, 23.7) |
| **Females** | | | | | | |
| All ages | 6.80 (6.20, 7.44) | 7.02 (6.41, 7.66) | 8.97 (8.29, 9.69) | 8.71 (8.04, 9.41) | 8.65 (8.00, 9.35) | 8.44 (7.79, 9.12) |
| 0–4 | 0.29 (0.01, 1.60) | 1.99 (0.80, 4.11) | 1.43 (0.46, 3.34) | 1.43 (0.46, 3.33) | 1.71 (0.63, 3.72) | 0.57 (0.07, 2.05) |
| 5–11 | 0.77 (0.21, 1.98) | 1.34 (0.54, 2.76) | 0.95 (0.31, 2.22) | 0.57 (0.12, 1.66) | 0.57 (0.12, 1.65) | 0.19 (0.00, 1.05) |
| 12–15 | 0.33 (0.01, 1.86) | 0.33 (0.01, 1.85) | 1.98 (0.73, 4.32) | 1.97 (0.72, 4.29) | 1.30 (0.35, 3.32) | 0.96 (0.20, 2.81) |
| 16–19 | 3.03 (1.45, 5.57) | 3.33 (1.66, 5.95) | 4.78 (2.73, 7.77) | 5.30 (3.14, 8.37) | 3.81 (2.03, 6.52) | 3.55 (1.83, 6.20) |
| 20–24 | 5.95 (3.92, 8.66) | 5.25 (3.37, 7.82) | 6.42 (4.33, 9.17) | 6.45 (4.38, 9.15) | 6.30 (4.28, 8.95) | 4.63 (2.94, 6.95) |
| 25–29 | 4.38 (2.68, 6.77) | 6.48 (4.38, 9.26) | 7.18 (4.97, 10.0) | 5.49 (3.62, 7.99) | 7.09 (4.96, 9.81) | 6.33 (4.36, 8.89) |
| 30–39 | 6.09 (4.60, 7.91) | 6.77 (5.20, 8.66) | 6.37 (4.86, 8.19) | 6.96 (5.40, 8.84) | 7.70 (6.06, 9.63) | 8.08 (6.43, 10.0) |
| 40–49 | 5.43 (4.05, 7.12) | 6.19 (4.72, 7.99) | 9.58 (7.71, 11.8) | 9.80 (7.91, 12.0) | 8.31 (6.58, 10.4) | 9.04 (7.23, 11.2) |
| 50–59 | 9.40 (7.63, 11.5) | 9.26 (7.51, 11.3) | 9.55 (7.77, 11.6) | 10.5 (8.67, 12.7) | 10.7 (8.79, 12.9) | 10.6 (8.68, 12.8) |
| 60–69 | 11.2 (9.02, 13.8) | 10.4 (8.31, 12.8) | 14.3 (11.9, 17.1) | 13.8 (11.4, 16.5) | 14.0 (11.6, 16.7) | 13.1 (10.8, 15.6) |
| 70–79 | 13.4 (10.3, 17.1) | 13.3 (10.3, 16.9) | 19.7 (16.1, 23.9) | 17.1 (13.8, 20.9) | 16.9 (13.7, 20.6) | 15.4 (12.4, 18.8) |
| ≥80 | 14.2 (10.5, 18.7) | 11.9 (8.62, 16.1) | 18.5 (14.3, 23.4) | 15.2 (11.5, 19.7) | 14.6 (11.0, 18.9) | 16.8 (13.0, 21.4) |
| **Males** | | | | | | |

| | | | | | | |
|---|---|---|---|---|---|---|
| All ages | 12.9 (12.1, 13.8) | 13.5 (12.7, 14.4) | 15.9 (15.0, 16.9) | 15.1 (14.2, 16.0) | 15.4 (14.5, 16.3) | 13.0 (12.2, 13.9) |
| 0–4 | 0.82 (0.17, 2.40) | 1.36 (0.44, 3.18) | 0.82 (0.17, 2.39) | 1.63 (0.60, 3.54) | 0.81 (0.17, 2.37) | 0.54 (0.07, 1.95) |
| 5–11 | 0.75 (0.20, 1.91) | 1.11 (0.41, 2.42) | 0.92 (0.30, 2.14) | 0.91 (0.30, 2.13) | 0.36 (0.04, 1.31) | 1.27 (0.51, 2.62) |
| 12–15 | 1.59 (0.52, 3.71) | 5.09 (2.91, 8.26) | 4.76 (2.66, 7.85) | 3.47 (1.73, 6.21) | 2.81 (1.28, 5.33) | 3.71 (1.91, 6.47) |
| 16–19 | 15.9 (12.0, 20.7) | 14.5 (10.8, 19.0) | 26.6 (21.5, 32.5) | 17.1 (13.1, 22.0) | 22.5 (17.9, 28.0) | 11.3 (8.04, 15.3) |
| 20–24 | 18.7 (15.1, 22.9) | 17.9 (14.4, 22.0) | 19.5 (15.8, 23.7) | 17.8 (14.4, 21.8) | 18.9 (15.4, 22.9) | 12.9 (10.1, 16.3) |
| 25–29 | 13.2 (10.1, 16.9) | 15.3 (12.0, 19.2) | 18.5 (14.9, 22.7) | 17.5 (14.1, 21.5) | 15.2 (12.1, 18.9) | 12.1 (9.34, 15.3) |
| 30–39 | 16.3 (13.8, 19.2) | 15.2 (12.7, 18.0) | 17.1 (14.6, 20.0) | 17.4 (14.9, 20.3) | 16.3 (13.8, 19.0) | 13.9 (11.7, 16.4) |
| 40–49 | 14.6 (12.2, 17.2) | 15.6 (13.2, 18.4) | 16.2 (13.7, 19.0) | 16.0 (13.5, 18.9) | 15.4 (12.9, 18.2) | 13.2 (10.9, 15.8) |
| 50–59 | 13.7 (11.5, 16.2) | 15.2 (12.9, 17.7) | 18.2 (15.7, 21.0) | 16.3 (14.0, 19.0) | 15.2 (12.9, 17.8) | 14.9 (12.6, 17.5) |
| 60–69 | 15.2 (12.5, 18.2) | 16.4 (13.7, 19.5) | 19.0 (16.0, 22.3) | 18.9 (16.0, 22.1) | 20.1 (17.2, 23.4) | 18.3 (15.5, 21.4) |
| 70–79 | 17.3 (13.6, 21.8) | 19.2 (15.3, 23.8) | 23.7 (19.5, 28.6) | 23.1 (19.0, 27.8) | 26.6 (22.3, 31.5) | 21.7 (18.0, 26.1) |
| ≥80 | 21.0 (15.4, 27.9) | 16.3 (11.6, 22.4) | 19.1 (14.0, 25.5) | 22.1 (16.7, 28.8) | 27.3 (21.3, 34.4) | 24.6 (19.0, 31.4) |
| **Myocarditis** | | | | | | |
| **Both sexes** | | | | | | |
| All ages | 2.54 (2.28, 2.82) | 2.88 (2.60, 3.17) | 3.38 (3.08, 3.69) | 2.58 (2.32, 2.86) | 2.93 (2.66, 3.22) | 2.42 (2.18, 2.69) |
| 0–4 | 0.56 (0.15, 1.44) | 1.25 (0.57, 2.38) | 0.70 (0.23, 1.63) | 1.11 (0.48, 2.19) | 0.83 (0.31, 1.81) | 0.55 (0.15, 1.42) |
| 5–11 | 0.38 (0.10, 0.97) | 0.94 (0.45, 1.73) | 0.28 (0.06, 0.82) | 0.46 (0.15, 1.08) | 0.19 (0.02, 0.67) | 0.19 (0.02, 0.67) |
| 12–15 | 0.16 (0.00, 0.91) | 1.30 (0.56, 2.56) | 0.97 (0.36, 2.12) | 0.64 (0.18, 1.65) | 0.64 (0.17, 1.63) | 1.10 (0.44, 2.27) |
| 16–19 | 4.25 (2.85, 6.11) | 4.10 (2.72, 5.93) | 7.52 (5.61, 9.86) | 4.28 (2.88, 6.10) | 5.56 (3.96, 7.61) | 3.46 (2.22, 5.15) |
| 20–24 | 5.71 (4.29, 7.45) | 4.09 (2.91, 5.59) | 4.61 (3.36, 6.17) | 3.97 (2.84, 5.41) | 4.26 (3.10, 5.72) | 2.89 (1.95, 4.12) |
| 25–29 | 2.81 (1.83, 4.11) | 4.97 (3.65, 6.61) | 4.43 (3.20, 5.96) | 3.96 (2.83, 5.39) | 4.01 (2.89, 5.42) | 2.32 (1.50, 3.43) |
| 30–39 | 4.12 (3.24, 5.17) | 3.67 (2.84, 4.66) | 3.60 (2.79, 4.58) | 3.14 (2.40, 4.05) | 3.20 (2.46, 4.09) | 3.44 (2.68, 4.35) |
| 40–49 | 2.55 (1.88, 3.38) | 3.10 (2.36, 4.01) | 4.73 (3.80, 5.83) | 2.86 (2.14, 3.74) | 3.40 (2.61, 4.35) | 2.43 (1.77, 3.25) |
| 50–59 | 1.98 (1.42, 2.68) | 2.98 (2.28, 3.82) | 3.41 (2.67, 4.30) | 2.12 (1.54, 2.85) | 2.62 (1.97, 3.42) | 2.45 (1.82, 3.23) |
| 60–69 | 2.00 (1.36, 2.84) | 2.57 (1.84, 3.48) | 2.82 (2.06, 3.76) | 2.39 (1.71, 3.26) | 3.32 (2.51, 4.30) | 2.49 (1.81, 3.35) |
| 70–79 | 2.12 (1.28, 3.32) | 2.16 (1.32, 3.33) | 2.94 (1.97, 4.22) | 2.98 (2.02, 4.23) | 3.03 (2.09, 4.26) | 3.17 (2.22, 4.39) |
| ≥80 | 2.94 (1.71, 4.71) | 1.68 (0.81, 3.10) | 3.28 (2.01, 5.07) | 2.24 (1.23, 3.76) | 2.96 (1.78, 4.63) | 3.05 (1.86, 4.71) |
| **Females** | | | | | | |
| All ages | 1.64 (1.35, 1.97) | 1.99 (1.67, 2.35) | 2.31 (1.97, 2.69) | 1.79 (1.50, 2.13) | 1.98 (1.67, 2.33) | 1.66 (1.38, 1.98) |
| 0–4 | 0.29 (0.01, 1.60) | 1.42 (0.46, 3.32) | 0.57 (0.07, 2.07) | 0.86 (0.18, 2.50) | 1.43 (0.46, 3.33) | 0.57 (0.07, 2.05) |
| 5–11 | 0.19 (0.00, 1.08) | 1.15 (0.42, 2.50) | 0.19 (0.00, 1.06) | 0.19 (0.00, 1.05) | 0.38 (0.05, 1.36) | 0.00 (0.00, 0.00) |
| 12–15 | 0.00 (0.00, 0.00) | 0.33 (0.01, 1.85) | 0.33 (0.01, 1.84) | 0.33 (0.01, 1.83) | 0.65 (0.08, 2.34) | 0.00 (0.00, 0.00) |
| 16–19 | 1.51 (0.49, 3.53) | 1.81 (0.67, 3.95) | 1.79 (0.66, 3.90) | 1.77 (0.65, 3.84) | 1.17 (0.32, 3.01) | 1.18 (0.32, 3.03) |
| 20–24 | 2.42 (1.21, 4.34) | 1.97 (0.90, 3.74) | 1.50 (0.60, 3.09) | 1.46 (0.59, 3.00) | 1.83 (0.84, 3.47) | 1.41 (0.57, 2.91) |
| 25–29 | 0.88 (0.24, 2.24) | 2.81 (1.50, 4.81) | 3.38 (1.93, 5.48) | 1.42 (0.57, 2.93) | 1.97 (0.94, 3.62) | 1.34 (0.54, 2.77) |

| | | | | | | |
|---|---|---|---|---|---|---|
| 30–39 | 2.07 (1.24, 3.23) | 2.36 (1.48, 3.58) | 1.80 (1.05, 2.89) | 1.56 (0.87, 2.57) | 1.01 (0.49, 1.86) | 2.17 (1.36, 3.28) |
| 40–49 | 1.25 (0.65, 2.19) | 1.99 (1.20, 3.11) | 3.05 (2.04, 4.38) | 1.69 (0.96, 2.74) | 2.63 (1.70, 3.88) | 1.89 (1.12, 2.99) |
| 50–59 | 2.30 (1.47, 3.42) | 2.20 (1.39, 3.29) | 2.39 (1.55, 3.53) | 2.01 (1.25, 3.08) | 2.21 (1.40, 3.32) | 2.04 (1.26, 3.11) |
| 60–69 | 2.24 (1.33, 3.55) | 2.42 (1.48, 3.73) | 3.66 (2.49, 5.20) | 3.00 (1.96, 4.40) | 3.04 (2.00, 4.42) | 2.41 (1.51, 3.65) |
| 70–79 | 2.50 (1.29, 4.37) | 2.02 (0.97, 3.71) | 3.23 (1.88, 5.16) | 3.60 (2.20, 5.56) | 2.75 (1.57, 4.47) | 1.98 (1.02, 3.46) |
| ≥80 | 1.98 (0.80, 4.08) | 1.66 (0.61, 3.62) | 3.53 (1.88, 6.03) | 1.86 (0.75, 3.84) | 3.38 (1.80, 5.78) | 2.30 (1.05, 4.36) |
| **Males** | | | | | | |
| All ages | 3.47 (3.04, 3.94) | 3.79 (3.34, 4.28) | 4.47 (3.99, 5.00) | 3.38 (2.97, 3.84) | 3.90 (3.45, 4.38) | 3.20 (2.80, 3.64) |
| 0–4 | 0.82 (0.17, 2.40) | 1.09 (0.30, 2.79) | 0.82 (0.17, 2.39) | 1.36 (0.44, 3.16) | 0.27 (0.01, 1.51) | 0.54 (0.07, 1.95) |
| 5–11 | 0.56 (0.12, 1.63) | 0.74 (0.20, 1.89) | 0.37 (0.04, 1.33) | 0.73 (0.20, 1.87) | 0.00 (0.00, 0.00) | 0.36 (0.04, 1.31) |
| 12–15 | 0.32 (0.01, 1.77) | 2.23 (0.89, 4.58) | 1.59 (0.52, 3.70) | 0.95 (0.20, 2.77) | 0.62 (0.08, 2.25) | 2.16 (0.87, 4.45) |
| 16–19 | 6.82 (4.37, 10.2) | 6.25 (3.91, 9.46) | 12.9 (9.43, 17.2) | 6.63 (4.25, 9.87) | 9.72 (6.77, 13.5) | 5.62 (3.44, 8.69) |
| 20–24 | 8.74 (6.32, 11.8) | 6.03 (4.07, 8.61) | 7.47 (5.28, 10.3) | 6.26 (4.31, 8.80) | 6.47 (4.51, 9.00) | 4.23 (2.68, 6.35) |
| 25–29 | 4.69 (2.94, 7.09) | 7.03 (4.87, 9.83) | 5.42 (3.57, 7.89) | 6.35 (4.37, 8.92) | 5.93 (4.06, 8.37) | 3.24 (1.92, 5.12) |
| 30–39 | 6.28 (4.73, 8.17) | 5.02 (3.66, 6.72) | 5.45 (4.05, 7.19) | 4.75 (3.47, 6.36) | 5.38 (4.03, 7.04) | 4.70 (3.47, 6.24) |
| 40–49 | 3.88 (2.72, 5.37) | 4.26 (3.03, 5.82) | 6.49 (4.94, 8.37) | 4.09 (2.88, 5.64) | 4.21 (2.98, 5.77) | 2.99 (1.97, 4.36) |
| 50–59 | 1.65 (0.96, 2.65) | 3.77 (2.68, 5.15) | 4.45 (3.26, 5.94) | 2.24 (1.42, 3.36) | 3.03 (2.06, 4.31) | 2.87 (1.92, 4.12) |
| 60–69 | 1.74 (0.93, 2.98) | 2.73 (1.69, 4.18) | 1.91 (1.07, 3.15) | 1.74 (0.95, 2.91) | 3.62 (2.44, 5.17) | 2.58 (1.62, 3.90) |
| 70–79 | 1.69 (0.68, 3.47) | 2.32 (1.11, 4.26) | 2.61 (1.35, 4.56) | 2.27 (1.13, 4.06) | 3.35 (1.95, 5.36) | 4.54 (2.91, 6.75) |
| ≥80 | 4.46 (2.14, 8.20) | 1.72 (0.47, 4.40) | 2.91 (1.17, 5.99) | 2.81 (1.13, 5.80) | 2.34 (0.86, 5.08) | 4.17 (2.08, 7.46) |
| **Pericarditis** | | | | | | |
| **Both sexes** | | | | | | |
| All ages | 7.53 (7.08, 8.00) | 7.64 (7.19, 8.11) | 9.35 (8.85, 9.87) | 9.44 (8.94, 9.96) | 9.24 (8.75, 9.75) | 8.44 (7.97, 8.92) |
| 0–4 | 0.00 (0.00, 0.00) | 0.42 (0.09, 1.22) | 0.56 (0.15, 1.43) | 0.42 (0.09, 1.22) | 0.42 (0.09, 1.22) | 0.00 (0.00, 0.00) |
| 5–11 | 0.57 (0.21, 1.24) | 0.38 (0.10, 0.96) | 0.65 (0.26, 1.35) | 0.28 (0.06, 0.81) | 0.28 (0.06, 0.81) | 0.56 (0.20, 1.21) |
| 12–15 | 0.98 (0.36, 2.13) | 1.79 (0.89, 3.20) | 2.59 (1.48, 4.21) | 2.25 (1.23, 3.78) | 1.43 (0.65, 2.72) | 1.26 (0.54, 2.48) |
| 16–19 | 6.16 (4.44, 8.32) | 5.86 (4.18, 7.98) | 9.98 (7.76, 12.6) | 7.70 (5.78, 10.0) | 8.85 (6.78, 11.3) | 4.32 (2.92, 6.17) |
| 20–24 | 7.50 (5.86, 9.47) | 8.70 (6.93, 10.8) | 9.22 (7.41, 11.3) | 8.73 (7.01, 10.8) | 9.00 (7.27, 11.0) | 6.35 (4.91, 8.08) |
| 25–29 | 6.37 (4.85, 8.22) | 6.34 (4.84, 8.16) | 8.95 (7.17, 11.0) | 8.11 (6.45, 10.1) | 7.73 (6.14, 9.61) | 7.15 (5.64, 8.93) |
| 30–39 | 7.46 (6.25, 8.84) | 7.39 (6.20, 8.75) | 8.50 (7.22, 9.93) | 9.32 (8.00, 10.8) | 8.98 (7.70, 10.4) | 7.81 (6.65, 9.13) |
| 40–49 | 7.48 (6.30, 8.82) | 8.03 (6.79, 9.42) | 8.39 (7.13, 9.82) | 10.1 (8.74, 11.7) | 8.52 (7.25, 9.96) | 8.74 (7.45, 10.2) |
| 50–59 | 9.70 (8.41, 11.1) | 9.51 (8.23, 10.9) | 10.6 (9.22, 12.1) | 11.4 (9.99, 12.9) | 10.4 (9.04, 11.9) | 10.5 (9.17, 12.0) |
| 60–69 | 11.2 (9.57, 13.0) | 10.9 (9.34, 12.6) | 13.9 (12.2, 15.8) | 13.8 (12.1, 15.7) | 13.6 (11.9, 15.5) | 13.3 (11.7, 15.1) |
| 70–79 | 13.2 (10.9, 15.8) | 14.0 (11.7, 16.6) | 18.7 (16.1, 21.7) | 16.9 (14.5, 19.6) | 18.6 (16.2, 21.4) | 15.3 (13.1, 17.8) |
| ≥80 | 14.0 (11.1, 17.4) | 12.1 (9.49, 15.3) | 15.8 (12.8, 19.3) | 15.7 (12.7, 19.1) | 16.7 (13.7, 20.2) | 16.9 (13.9, 20.4) |
| **Females** | | | | | | |

| | | | | | | |
|---|---|---|---|---|---|---|
| All ages | 5.24 (4.72, 5.81) | 5.20 (4.68, 5.76) | 6.81 (6.22, 7.45) | 7.00 (6.40, 7.63) | 6.70 (6.12, 7.32) | 6.88 (6.30, 7.50) |
| 0–4 | 0.00 (0.00, 0.00) | 0.57 (0.07, 2.06) | 0.86 (0.18, 2.51) | 0.57 (0.07, 2.06) | 0.29 (0.01, 1.59) | 0.00 (0.00, 0.00) |
| 5–11 | 0.58 (0.12, 1.70) | 0.38 (0.05, 1.38) | 0.76 (0.21, 1.95) | 0.38 (0.05, 1.37) | 0.19 (0.00, 1.05) | 0.19 (0.00, 1.05) |
| 12–15 | 0.33 (0.01, 1.86) | 0.33 (0.01, 1.85) | 1.65 (0.54, 3.86) | 1.64 (0.53, 3.83) | 0.65 (0.08, 2.34) | 0.96 (0.20, 2.81) |
| 16–19 | 1.82 (0.67, 3.95) | 2.12 (0.85, 4.36) | 3.29 (1.64, 5.88) | 3.53 (1.82, 6.17) | 2.64 (1.21, 5.01) | 2.37 (1.02, 4.66) |
| 20–24 | 3.75 (2.18, 6.00) | 3.72 (2.17, 5.96) | 4.92 (3.12, 7.39) | 5.20 (3.37, 7.68) | 4.47 (2.80, 6.77) | 3.42 (1.99, 5.48) |
| 25–29 | 3.50 (2.00, 5.69) | 3.67 (2.14, 5.88) | 4.01 (2.41, 6.26) | 4.48 (2.81, 6.78) | 5.12 (3.34, 7.50) | 4.99 (3.26, 7.31) |
| 30–39 | 4.24 (3.02, 5.80) | 4.41 (3.16, 5.98) | 4.67 (3.39, 6.27) | 5.51 (4.13, 7.21) | 6.68 (5.17, 8.50) | 6.01 (4.60, 7.73) |
| 40–49 | 4.18 (2.98, 5.69) | 4.30 (3.09, 5.84) | 6.63 (5.10, 8.49) | 8.22 (6.50, 10.3) | 5.68 (4.27, 7.41) | 7.14 (5.55, 9.06) |
| 50–59 | 7.10 (5.57, 8.91) | 7.35 (5.80, 9.19) | 7.36 (5.80, 9.19) | 8.63 (6.94, 10.6) | 8.57 (6.88, 10.6) | 8.83 (7.11, 10.8) |
| 60–69 | 9.10 (7.13, 11.4) | 8.09 (6.27, 10.3) | 10.9 (8.76, 13.3) | 10.8 (8.67, 13.2) | 10.9 (8.86, 13.3) | 10.9 (8.83, 13.2) |
| 70–79 | 10.9 (8.10, 14.2) | 11.5 (8.70, 14.9) | 16.7 (13.4, 20.6) | 13.5 (10.6, 16.9) | 14.3 (11.4, 17.7) | 13.5 (10.8, 16.8) |
| ≥80 | 12.5 (9.05, 16.7) | 10.3 (7.22, 14.1) | 15.5 (11.7, 20.1) | 13.3 (9.87, 17.5) | 11.2 (8.09, 15.1) | 14.5 (11.0, 18.8) |
| **Males** | | | | | | |
| All ages | 9.89 (9.15, 10. 7) | 10.2 (9.41, 10.9) | 12.0 (11.2, 12.8) | 12.0 (11.2, 12.8) | 11.8 (11.1, 12.7) | 10.0 (9.31, 10.8) |
| 0–4 | 0.00 (0.00, 0.00) | 0.27 (0.01, 1.52) | 0.27 (0.01, 1.52) | 0.27 (0.01, 1.51) | 0.54 (0.07, 1.95) | 0.00 (0.00, 0.00) |
| 5–11 | 0.56 (0.12, 1.63) | 0.37 (0.04, 1.34) | 0.55 (0.11, 1.61) | 0.18 (0.00, 1.02) | 0.36 (0.04, 1.31) | 0.91 (0.30, 2.12) |
| 12–15 | 1.59 (0.52, 3.71) | 3.18 (1.52, 5.85) | 3.49 (1.74, 6.25) | 2.84 (1.30, 5.39) | 2.18 (0.88, 4.50) | 1.54 (0.50, 3.60) |
| 16–19 | 10.2 (7.17, 14.2) | 9.37 (6.45, 13.2) | 16.2 (12.3, 21.0) | 11.6 (8.37, 15.7) | 14.7 (11.0, 19.3) | 6.19 (3.88, 9.37) |
| 20–24 | 11.0 (8.24, 14.3) | 13.3 (10.3, 16.9) | 13.2 (10.2, 16.7) | 12.0 (9.19, 15.3) | 13.1 (10.3, 16.6) | 9.02 (6.67, 11.9) |
| 25–29 | 9.16 (6.63, 12.3) | 8.89 (6.44, 12.0) | 13.7 (10.6, 17.3) | 11.6 (8.81, 14.9) | 10.2 (7.68, 13.3) | 9.17 (6.83, 12.1) |
| 30–39 | 10.8 (8.77, 13.3) | 10.5 (8.48, 12.8) | 12.4 (10.3, 14.9) | 13.2 (11.0, 15.7) | 11.3 (9.28, 13.6) | 9.60 (7.80, 11.7) |
| 40–49 | 10.9 (8.87, 13.2) | 11.9 (9.77, 14.4) | 10.2 (8.26, 12.5) | 12.2 (9.99, 14.7) | 11.5 (9.41, 14.0) | 10.4 (8.43, 12.8) |
| 50–59 | 12.3 (10.3, 14.7) | 11.7 (9.70, 14.0) | 13.8 (11.7, 16.3) | 14.2 (12.0, 16.7) | 12.2 (10.2, 14.6) | 12.3 (10.2, 14.6) |
| 60–69 | 13.4 (10.9, 16.3) | 13.9 (11.4, 16.8) | 17.2 (14.4, 20.3) | 17.1 (14.4, 20.2) | 16.5 (13.9, 19.5) | 15.9 (13.4, 18.9) |
| 70–79 | 15.9 (12.3, 20.2) | 16.9 (13.3, 21.3) | 21.1 (17.1, 25.7) | 20.8 (17.0, 25.3) | 23.6 (19.6, 28.3) | 17.4 (14.0, 21.3) |
| ≥80 | 16.5 (11.6, 22.7) | 15.0 (10.5, 20.9) | 16.2 (11.5, 22.2) | 19.3 (14.2, 25.6) | 24.9 (19.2, 31.8) | 20.5 (15.4, 26.7) |

Supplemental Table S7: Annual background rates of hospitalizations and emergency department visits for Kawasaki disease by age group and sex in Ontario, 2015 to 2020

| Age group, years | Incidence rate per 100,000 population (95% confidence interval) | | | | | |
|---|---|---|---|---|---|---|
| | 2015 | 2016 | 2017 | 2018 | 2019 | 2020 |
| **Both sexes** | | | | | | |
| All ages | 1.96 (1.73, 2.21) | 2.00 (1.77, 2.25) | 2.03 (1.80, 2.27) | 2.15 (1.92, 2.41) | 1.71 (1.51, 1.94) | 1.55 (1.35, 1.76) |
| 0–4 | 25.2 (21.7, 29.2) | 26.3 (22.7, 30.4) | 25.9 (22.4, 30.0) | 28.9 (25.1, 33.1) | 23.2 (19.8, 27.0) | 21.4 (18.2, 25.1) |
| 5–11 | 7.50 (5.94, 9.35) | 7.34 (5.80, 9.16) | 7.48 (5.93, 9.31) | 8.08 (6.47, 9.97) | 6.20 (4.81, 7.88) | 5.28 (4.00, 6.85) |
| 12–15 | 0.82 (0.26, 1.90) | 0.97 (0.36, 2.12) | 1.94 (1.00, 3.39) | 0.80 (0.26, 1.88) | 1.43 (0.65, 2.72) | 1.89 (0.98, 3.30) |
| 16–19 | 0.44 (0.09, 1.29) | 0.00 (0.00, 0.00) | 0.58 (0.16, 1.48) | 0.14 (0.00, 0.79) | 0.14 (0.00, 0.79) | 0.14 (0.00, 0.80) |
| 20–24 | 0.00 (0.00, 0.00) | 0.10 (0.00, 0.58) | 0.10 (0.00, 0.57) | 0.40 (0.11, 1.02) | 0.29 (0.06, 0.85) | 0.00 (0.00, 0.00) |
| 25–29 | 0.00 (0.00, 0.00) | 0.00 (0.00, 0.00) | 0.00 (0.00, 0.00) | 0.10 (0.00, 0.55) | 0.10 (0.00, 0.53) | 0.00 (0.00, 0.00) |
| 30–39 | 0.00 (0.00, 0.00) | 0.00 (0.00, 0.00) | 0.11 (0.01, 0.39) | 0.10 (0.01, 0.38) | 0.00 (0.00, 0.00) | 0.00 (0.00, 0.00) |
| 40–49 | 0.05 (0.00, 0.30) | 0.05 (0.00, 0.30) | 0.00 (0.00, 0.00) | 0.00 (0.00, 0.00) | 0.00 (0.00, 0.00) | 0.00 (0.00, 0.00) |
| 50–59 | 0.00 (0.00, 0.00) | 0.05 (0.00, 0.27) | 0.00 (0.00, 0.00) | 0.00 (0.00, 0.00) | 0.05 (0.00, 0.27) | 0.05 (0.00, 0.27) |
| 60–69 | 0.06 (0.00, 0.36) | 0.06 (0.00, 0.35) | 0.00 (0.00, 0.00) | 0.00 (0.00, 0.00) | 0.00 (0.00, 0.00) | 0.00 (0.00, 0.00) |
| 70–79 | 0.00 (0.00, 0.00) | 0.00 (0.00, 0.00) | 0.00 (0.00, 0.00) | 0.00 (0.00, 0.00) | 0.00 (0.00, 0.00) | 0.18 (0.02, 0.64) |
| ≥80 | 0.00 (0.00, 0.00) | 0.00 (0.00, 0.00) | 0.00 (0.00, 0.00) | 0.00 (0.00, 0.00) | 0.00 (0.00, 0.00) | 0.00 (0.00, 0.00) |
| **Females** | | | | | | |
| All ages | 1.68 (1.39, 2.01) | 1.58 (1.30, 1.90) | 1.39 (1.13, 1.69) | 1.71 (1.42, 2.04) | 1.51 (1.24, 1.82) | 1.26 (1.02, 1.54) |
| 0–4 | 24.4 (19.5, 30.1) | 22.8 (18.1, 28.4) | 17.4 (13.3, 22.4) | 21.1 (16.6, 26.5) | 23.9 (19.1, 29.7) | 18.2 (14.0, 23.2) |
| 5–11 | 5.23 (3.44, 7.60) | 5.56 (3.72, 7.98) | 6.10 (4.17, 8.61) | 8.71 (6.37, 11.6) | 4.34 (2.75, 6.51) | 5.11 (3.37, 7.43) |
| 12–15 | 1.00 (0.21, 2.93) | 0.33 (0.01, 1.85) | 0.99 (0.20, 2.90) | 0.33 (0.01, 1.83) | 0.32 (0.01, 1.81) | 0.64 (0.08, 2.32) |
| 16–19 | 0.00 (0.00, 0.00) | 0.00 (0.00, 0.00) | 0.90 (0.18, 2.62) | 0.00 (0.00, 0.00) | 0.00 (0.00, 0.00) | 0.00 (0.00, 0.00) |
| 20–24 | 0.00 (0.00, 0.00) | 0.00 (0.00, 0.00) | 0.00 (0.00, 0.00) | 0.62 (0.13, 1.82) | 0.20 (0.01, 1.13) | 0.00 (0.00, 0.00) |
| 25–29 | 0.00 (0.00, 0.00) | 0.00 (0.00, 0.00) | 0.00 (0.00, 0.00) | 0.00 (0.00, 0.00) | 0.20 (0.00, 1.10) | 0.00 (0.00, 0.00) |
| 30–39 | 0.00 (0.00, 0.00) | 0.00 (0.00, 0.00) | 0.00 (0.00, 0.00) | 0.00 (0.00, 0.00) | 0.00 (0.00, 0.00) | 0.00 (0.00, 0.00) |
| 40–49 | 0.10 (0.00, 0.58) | 0.00 (0.00, 0.00) | 0.00 (0.00, 0.00) | 0.00 (0.00, 0.00) | 0.00 (0.00, 0.00) | 0.00 (0.00, 0.00) |
| 50–59 | 0.00 (0.00, 0.00) | 0.10 (0.00, 0.53) | 0.00 (0.00, 0.00) | 0.00 (0.00, 0.00) | 0.10 (0.00, 0.54) | 0.00 (0.00, 0.00) |
| 60–69 | 0.12 (0.00, 0.69) | 0.00 (0.00, 0.00) | 0.00 (0.00, 0.00) | 0.00 (0.00, 0.00) | 0.00 (0.00, 0.00) | 0.00 (0.00, 0.00) |
| 70–79 | 0.00 (0.00, 0.00) | 0.00 (0.00, 0.00) | 0.00 (0.00, 0.00) | 0.00 (0.00, 0.00) | 0.00 (0.00, 0.00) | 0.17 (0.00, 0.92) |
| ≥80 | 0.00 (0.00, 0.00) | 0.00 (0.00, 0.00) | 0.00 (0.00, 0.00) | 0.00 (0.00, 0.00) | 0.00 (0.00, 0.00) | 0.00 (0.00, 0.00) |
| **Males** | | | | | | |
| All ages | 2.25 (1.91, 2.64) | 2.43 (2.07, 2.83) | 2.68 (2.31, 3.10) | 2.61 (2.24, 3.01) | 1.92 (1.61, 2.27) | 1.84 (1.54, 2.18) |

| | | | | | | |
|---|---|---|---|---|---|---|
| 0–4 | 9.69 (7.24, 12.7) | 9.06 (6.70, 12.0) | 8.82 (6.50, 11.7) | 7.48 (5.37, 10.1) | 8.00 (5.81, 10.7) | 5.45 (3.68, 7.79) |
| 5–11 | 0.64 (0.08, 2.30) | 1.59 (0.52, 3.71) | 2.86 (1.31, 5.42) | 1.26 (0.34, 3.23) | 2.50 (1.08, 4.92) | 3.09 (1.48, 5.68) |
| 12–15 | 0.85 (0.18, 2.49) | 0.00 (0.00, 0.00) | 0.28 (0.01, 1.56) | 0.28 (0.01, 1.54) | 0.28 (0.01, 1.55) | 0.28 (0.01, 1.57) |
| 16–19 | 0.00 (0.00, 0.00) | 0.20 (0.01, 1.12) | 0.20 (0.00, 1.09) | 0.19 (0.00, 1.06) | 0.37 (0.04, 1.34) | 0.00 (0.00, 0.00) |
| 20–24 | 0.00 (0.00, 0.00) | 0.00 (0.00, 0.00) | 0.00 (0.00, 0.00) | 0.19 (0.00, 1.07) | 0.00 (0.00, 0.00) | 0.00 (0.00, 0.00) |
| 25–29 | 0.00 (0.00, 0.00) | 0.00 (0.00, 0.00) | 0.22 (0.03, 0.79) | 0.21 (0.03, 0.76) | 0.00 (0.00, 0.00) | 0.00 (0.00, 0.00) |
| 30–39 | 0.00 (0.00, 0.00) | 0.11 (0.00, 0.61) | 0.00 (0.00, 0.00) | 0.00 (0.00, 0.00) | 0.00 (0.00, 0.00) | 0.00 (0.00, 0.00) |
| 40–49 | 0.00 (0.00, 0.00) | 0.00 (0.00, 0.00) | 0.00 (0.00, 0.00) | 0.00 (0.00, 0.00) | 0.00 (0.00, 0.00) | 0.10 (0.00, 0.55) |
| 50–59 | 0.00 (0.00, 0.00) | 0.13 (0.00, 0.73) | 0.00 (0.00, 0.00) | 0.00 (0.00, 0.00) | 0.00 (0.00, 0.00) | 0.00 (0.00, 0.00) |
| 60–69 | 0.00 (0.00, 0.00) | 0.00 (0.00, 0.00) | 0.00 (0.00, 0.00) | 0.00 (0.00, 0.00) | 0.00 (0.00, 0.00) | 0.19 (0.00, 1.05) |
| 70–79 | 0.00 (0.00, 0.00) | 0.00 (0.00, 0.00) | 0.00 (0.00, 0.00) | 0.00 (0.00, 0.00) | 0.00 (0.00, 0.00) | 0.00 (0.00, 0.00) |
| ≥80 | 9.69 (7.24, 12.7) | 9.06 (6.70, 12.0) | 8.82 (6.50, 11.7) | 7.48 (5.37, 10.1) | 8.00 (5.81, 10.73) | 5.45 (3.68, 7.79) |

Supplemental Table S8: Annual background rates of hospitalizations and emergency department visits for Guillain-Barré Syndrome by age group and sex in Ontario, 2015 to 2020

| Age group, years | Incidence rate per 100,000 population (95% confidence interval) | | | | | |
|---|---|---|---|---|---|---|
| | **2015** | **2016** | **2017** | **2018** | **2019** | **2020** |
| **Both sexes** | | | | | | |
| All ages | 1.83 (1.61, 2.07) | 1.94 (1.71, 2.18) | 1.83 (1.61, 2.06) | 1.88 (1.66, 2.12) | 1.82 (1.60, 2.05) | 1.34 (1.16, 1.54) |
| 0–4 | 0.84 (0.31, 1.83) | 0.56 (0.15, 1.43) | 1.12 (0.48, 2.20) | 0.83 (0.31, 1.82) | 0.28 (0.03, 1.00) | 0.83 (0.30, 1.81) |
| 5–11 | 0.66 (0.27, 1.37) | 0.94 (0.45, 1.73) | 0.47 (0.15, 1.09) | 0.46 (0.15, 1.08) | 0.28 (0.06, 0.81) | 0.19 (0.02, 0.67) |
| 12–15 | 0.82 (0.26, 1.90) | 0.49 (0.10, 1.42) | 0.16 (0.00, 0.90) | 0.64 (0.18, 1.65) | 0.48 (0.10, 1.39) | 0.16 (0.00, 0.88) |
| 16–19 | 0.88 (0.32, 1.91) | 1.03 (0.41, 2.11) | 1.01 (0.41, 2.09) | 1.14 (0.49, 2.25) | 0.86 (0.31, 1.86) | 0.29 (0.03, 1.04) |
| 20–24 | 0.63 (0.23, 1.38) | 1.05 (0.50, 1.93) | 1.13 (0.56, 2.02) | 0.79 (0.34, 1.56) | 0.77 (0.33, 1.53) | 0.48 (0.16, 1.12) |
| 25–29 | 1.40 (0.75, 2.40) | 1.27 (0.66, 2.22) | 0.93 (0.42, 1.76) | 1.29 (0.68, 2.20) | 1.05 (0.52, 1.88) | 0.93 (0.45, 1.71) |
| 30–39 | 1.50 (0.99, 2.19) | 1.92 (1.33, 2.67) | 1.45 (0.96, 2.11) | 1.73 (1.19, 2.43) | 1.78 (1.24, 2.47) | 0.79 (0.45, 1.28) |
| 40–49 | 1.70 (1.16, 2.40) | 1.45 (0.95, 2.10) | 1.56 (1.04, 2.24) | 1.35 (0.87, 1.99) | 1.78 (1.23, 2.50) | 1.51 (1.00, 2.18) |
| 50–59 | 1.79 (1.26, 2.46) | 2.45 (1.82, 3.22) | 2.69 (2.03, 3.50) | 2.08 (1.50, 2.80) | 2.28 (1.68, 3.03) | 1.57 (1.07, 2.21) |
| 60–69 | 3.49 (2.62, 4.55) | 3.57 (2.70, 4.63) | 3.19 (2.38, 4.18) | 3.53 (2.69, 4.55) | 3.73 (2.87, 4.76) | 2.32 (1.67, 3.15) |
| 70–79 | 4.14 (2.91, 5.70) | 4.31 (3.08, 5.87) | 3.44 (2.39, 4.81) | 3.84 (2.75, 5.24) | 3.21 (2.24, 4.47) | 3.61 (2.59, 4.90) |
| ≥80 | 3.64 (2.25, 5.56) | 2.19 (1.17, 3.74) | 2.96 (1.75, 4.67) | 4.00 (2.59, 5.91) | 2.65 (1.54, 4.24) | 1.98 (1.06, 3.39) |
| **Females** | | | | | | |
| All ages | 1.59 (1.31, 1.92) | 1.73 (1.44, 2.07) | 1.44 (1.18, 1.75) | 1.46 (1.20, 1.77) | 1.64 (1.36, 1.96) | 1.06 (0.84, 1.32) |
| 0–4 | 0.86 (0.18, 2.51) | 0.57 (0.07, 2.06) | 0.57 (0.07, 2.07) | 0.86 (0.18, 2.50) | 0.29 (0.01, 1.59) | 1.14 (0.31, 2.91) |
| 5–11 | 0.97 (0.31, 2.26) | 0.77 (0.21, 1.96) | 0.38 (0.05, 1.38) | 0.00 (0.00, 0.00) | 0.19 (0.00, 1.05) | 0.38 (0.05, 1.37) |
| 12–15 | 0.33 (0.01, 1.86) | 0.66 (0.08, 2.40) | 0.00 (0.00, 0.00) | 0.66 (0.08, 2.37) | 0.00 (0.00, 0.00) | 0.00 (0.00, 0.00) |
| 16–19 | 0.61 (0.07, 2.19) | 0.91 (0.19, 2.65) | 0.60 (0.07, 2.16) | 0.59 (0.07, 2.13) | 0.59 (0.07, 2.12) | 0.30 (0.01, 1.65) |
| 20–24 | 0.66 (0.14, 1.93) | 1.31 (0.48, 2.86) | 1.28 (0.47, 2.80) | 0.42 (0.05, 1.50) | 1.02 (0.33, 2.37) | 0.81 (0.22, 2.06) |
| 25–29 | 0.88 (0.24, 2.24) | 1.08 (0.35, 2.52) | 0.63 (0.13, 1.85) | 1.63 (0.70, 3.21) | 0.59 (0.12, 1.73) | 1.15 (0.42, 2.50) |
| 30–39 | 1.09 (0.52, 2.00) | 1.40 (0.74, 2.39) | 1.06 (0.51, 1.95) | 0.73 (0.29, 1.50) | 1.82 (1.08, 2.88) | 0.69 (0.28, 1.42) |
| 40–49 | 1.57 (0.88, 2.58) | 1.15 (0.58, 2.07) | 0.95 (0.43, 1.80) | 1.26 (0.65, 2.21) | 1.58 (0.88, 2.60) | 1.26 (0.65, 2.20) |
| 50–59 | 1.63 (0.95, 2.61) | 2.58 (1.70, 3.75) | 2.87 (1.93, 4.09) | 1.92 (1.17, 2.96) | 1.93 (1.18, 2.97) | 0.97 (0.47, 1.78) |
| 60–69 | 2.62 (1.62, 4.00) | 3.02 (1.95, 4.46) | 2.01 (1.17, 3.22) | 3.00 (1.96, 4.40) | 4.17 (2.93, 5.74) | 1.10 (0.53, 2.02) |
| 70–79 | 3.13 (1.75, 5.16) | 3.63 (2.15, 5.74) | 2.66 (1.45, 4.46) | 2.70 (1.51, 4.45) | 2.41 (1.32, 4.04) | 3.30 (2.02, 5.10) |
| ≥80 | 4.25 (2.38, 7.01) | 1.66 (0.61, 3.62) | 2.17 (0.94, 4.28) | 2.39 (1.09, 4.55) | 1.30 (0.42, 3.03) | 0.77 (0.16, 2.24) |
| **Males** | | | | | | |
| All ages | 2.08 (1.75, 2.45) | 2.15 (1.82, 2.53) | 2.22 (1.88, 2.60) | 2.31 (1.97, 2.69) | 1.99 (1.68, 2.34) | 1.62 (1.34, 1.94) |

| | | | | | | |
|---|---|---|---|---|---|---|
| 0–4 | 0.82 (0.17, 2.40) | 0.54 (0.07, 1.97) | 1.63 (0.60, 3.56) | 0.81 (0.17, 2.38) | 0.27 (0.01, 1.51) | 0.54 (0.07, 1.95) |
| 5–11 | 0.37 (0.05, 1.35) | 1.11 (0.41, 2.42) | 0.55 (0.11, 1.61) | 0.91 (0.30, 2.13) | 0.36 (0.04, 1.31) | 0.00 (0.00, 0.00) |
| 12–15 | 1.27 (0.35, 3.26) | 0.32 (0.01, 1.77) | 0.32 (0.01, 1.77) | 0.63 (0.08, 2.28) | 0.94 (0.19, 2.74) | 0.31 (0.01, 1.72) |
| 16–19 | 1.14 (0.31, 2.91) | 1.14 (0.31, 2.91) | 1.40 (0.45, 3.27) | 1.66 (0.61, 3.61) | 1.11 (0.30, 2.84) | 0.28 (0.01, 1.57) |
| 20–24 | 0.61 (0.13, 1.78) | 0.80 (0.22, 2.06) | 0.98 (0.32, 2.29) | 1.14 (0.42, 2.48) | 0.55 (0.11, 1.62) | 0.18 (0.00, 1.03) |
| 25–29 | 1.92 (0.88, 3.64) | 1.45 (0.58, 2.98) | 1.21 (0.44, 2.62) | 0.96 (0.31, 2.25) | 1.48 (0.64, 2.92) | 0.72 (0.20, 1.84) |
| 30–39 | 1.94 (1.13, 3.11) | 2.45 (1.54, 3.72) | 1.85 (1.08, 2.97) | 2.75 (1.79, 4.02) | 1.73 (1.01, 2.77) | 0.88 (0.40, 1.67) |
| 40–49 | 1.83 (1.07, 2.94) | 1.75 (1.00, 2.84) | 2.20 (1.34, 3.40) | 1.44 (0.76, 2.46) | 1.99 (1.18, 3.15) | 1.77 (1.01, 2.88) |
| 50–59 | 1.94 (1.19, 3.00) | 2.32 (1.49, 3.45) | 2.52 (1.64, 3.69) | 2.24 (1.42, 3.36) | 2.64 (1.74, 3.84) | 2.18 (1.36, 3.29) |
| 60–69 | 4.43 (3.05, 6.22) | 4.16 (2.85, 5.88) | 4.45 (3.10, 6.19) | 4.09 (2.82, 5.75) | 3.26 (2.15, 4.74) | 3.63 (2.47, 5.16) |
| 70–79 | 5.30 (3.32, 8.02) | 5.09 (3.19, 7.71) | 4.35 (2.66, 6.72) | 5.15 (3.34, 7.61) | 4.14 (2.56, 6.32) | 3.97 (2.46, 6.07) |
| ≥80 | 2.67 (0.98, 5.82) | 3.01 (1.21, 6.19) | 4.16 (1.99, 7.64) | 6.43 (3.68, 10.45) | 4.67 (2.41, 8.16) | 3.79 (1.82, 6.97) |

Supplemental Table S9: Annual background rates of hospitalizations and emergency department visits for transverse myelitis by age group and sex in Ontario, 2015 to 2020

| Age group, years | Incidence rate per 100,000 population (95% confidence interval) | | | | | |
|---|---|---|---|---|---|---|
| | **2015** | **2016** | **2017** | **2018** | **2019** | **2020** |
| **Transverse myelitis, narrow definition** | | | | | | |
| **Both sexes** | | | | | | |
| All ages | 0.61 (0.49, 0.76) | 0.90 (0.75, 1.07) | 0.73 (0.60, 0.89) | 0.85 (0.71, 1.02) | 0.87 (0.73, 1.04) | 0.83 (0.69, 1.00) |
| 0–4 | 0.28 (0.03, 1.01) | 0.28 (0.03, 1.01) | 0.28 (0.03, 1.01) | 0.56 (0.15, 1.42) | 0.28 (0.03, 1.00) | 0.14 (0.00, 0.77) |
| 5–11 | 0.28 (0.06, 0.83) | 0.19 (0.02, 0.68) | 0.09 (0.00, 0.52) | 0.28 (0.06, 0.81) | 0.46 (0.15, 1.08) | 0.19 (0.02, 0.67) |
| 12–15 | 0.16 (0.00, 0.91) | 0.65 (0.18, 1.66) | 0.49 (0.10, 1.42) | 0.00 (0.00, 0.00) | 0.00 (0.00, 0.00) | 0.16 (0.00, 0.88) |
| 16–19 | 0.29 (0.04, 1.06) | 0.44 (0.09, 1.28) | 0.72 (0.23, 1.69) | 0.86 (0.31, 1.86) | 1.00 (0.40, 2.06) | 0.29 (0.03, 1.04) |
| 20–24 | 0.21 (0.03, 0.76) | 0.63 (0.23, 1.37) | 0.20 (0.02, 0.74) | 0.79 (0.34, 1.56) | 0.48 (0.16, 1.13) | 0.48 (0.16, 1.12) |
| 25–29 | 0.22 (0.03, 0.78) | 1.27 (0.66, 2.22) | 0.93 (0.42, 1.76) | 0.89 (0.41, 1.69) | 0.29 (0.06, 0.84) | 0.65 (0.26, 1.34) |
| 30–39 | 0.67 (0.35, 1.17) | 1.15 (0.71, 1.76) | 0.48 (0.22, 0.92) | 0.73 (0.40, 1.23) | 0.96 (0.58, 1.50) | 1.23 (0.80, 1.81) |
| 40–49 | 1.01 (0.61, 1.57) | 0.86 (0.49, 1.39) | 1.02 (0.62, 1.60) | 1.35 (0.87, 1.99) | 0.81 (0.45, 1.33) | 0.97 (0.58, 1.53) |
| 50–59 | 0.72 (0.41, 1.19) | 1.20 (0.78, 1.77) | 1.20 (0.78, 1.77) | 1.01 (0.63, 1.55) | 0.92 (0.56, 1.44) | 0.93 (0.56, 1.45) |
| 60–69 | 0.65 (0.31, 1.19) | 0.75 (0.39, 1.31) | 0.86 (0.47, 1.44) | 1.02 (0.59, 1.63) | 1.05 (0.62, 1.66) | 0.96 (0.56, 1.54) |
| 70–79 | 0.78 (0.31, 1.61) | 1.83 (1.07, 2.93) | 0.91 (0.42, 1.73) | 0.87 (0.40, 1.64) | 2.20 (1.41, 3.28) | 1.06 (0.55, 1.85) |
| ≥80 | 1.56 (0.71, 2.96) | 0.84 (0.27, 1.97) | 0.82 (0.27, 1.92) | 0.96 (0.35, 2.09) | 1.56 (0.75, 2.87) | 2.13 (1.17, 3.58) |
| **Females** | | | | | | |
| All ages | 0.76 (0.57, 1.00) | 1.18 (0.94, 1.46) | 0.93 (0.72, 1.18) | 1.15 (0.91, 1.42) | 0.94 (0.73, 1.19) | 0.89 (0.68, 1.13) |
| 0–4 | 0.29 (0.01, 1.60) | 0.00 (0.00, 0.00) | 0.29 (0.01, 1.59) | 0.57 (0.07, 2.06) | 0.00 (0.00, 0.00) | 0.28 (0.01, 1.58) |
| 5–11 | 0.19 (0.00, 1.08) | 0.19 (0.00, 1.07) | 0.00 (0.00, 0.00) | 0.38 (0.05, 1.37) | 0.00 (0.00, 0.00) | 0.00 (0.00, 0.00) |
| 12–15 | 0.00 (0.00, 0.00) | 1.00 (0.21, 2.91) | 0.99 (0.20, 2.90) | 0.00 (0.00, 0.00) | 0.00 (0.00, 0.00) | 0.00 (0.00, 0.00) |
| 16–19 | 0.61 (0.07, 2.19) | 0.60 (0.07, 2.18) | 0.90 (0.18, 2.62) | 0.88 (0.18, 2.58) | 2.05 (0.83, 4.23) | 0.59 (0.07, 2.14) |
| 20–24 | 0.22 (0.01, 1.23) | 0.66 (0.14, 1.92) | 0.21 (0.01, 1.19) | 1.66 (0.72, 3.28) | 0.61 (0.13, 1.78) | 0.40 (0.05, 1.46) |
| 25–29 | 0.44 (0.05, 1.58) | 2.16 (1.04, 3.98) | 1.48 (0.59, 3.04) | 1.42 (0.57, 2.93) | 0.59 (0.12, 1.73) | 0.96 (0.31, 2.24) |
| 30–39 | 0.87 (0.38, 1.71) | 1.29 (0.67, 2.25) | 0.64 (0.23, 1.39) | 0.94 (0.43, 1.78) | 0.81 (0.35, 1.60) | 1.08 (0.54, 1.94) |
| 40–49 | 1.67 (0.95, 2.71) | 1.26 (0.65, 2.20) | 1.47 (0.81, 2.47) | 2.00 (1.21, 3.13) | 1.05 (0.50, 1.93) | 1.58 (0.88, 2.60) |
| 50–59 | 0.77 (0.33, 1.51) | 1.53 (0.87, 2.48) | 1.43 (0.80, 2.36) | 1.34 (0.73, 2.25) | 1.06 (0.53, 1.90) | 0.87 (0.40, 1.66) |
| 60–69 | 0.62 (0.20, 1.45) | 1.21 (0.58, 2.22) | 1.18 (0.57, 2.17) | 1.04 (0.48, 1.97) | 1.01 (0.46, 1.92) | 0.66 (0.24, 1.43) |
| 70–79 | 1.25 (0.46, 2.72) | 2.02 (0.97, 3.71) | 0.76 (0.21, 1.94) | 0.90 (0.29, 2.10) | 1.89 (0.94, 3.39) | 1.16 (0.46, 2.38) |
| ≥80 | 0.85 (0.18, 2.48) | 1.11 (0.30, 2.84) | 0.54 (0.07, 1.96) | 1.33 (0.43, 3.10) | 1.82 (0.73, 3.75) | 2.04 (0.88, 4.02) |
| **Males** | | | | | | |

| | | | | | | |
|---|---|---|---|---|---|---|
| All ages | 0.46 (0.31, 0.65) | 0.61 (0.44, 0.83) | 0.53 (0.38, 0.74) | 0.55 (0.39, 0.75) | 0.81 (0.61, 1.04) | 0.78 (0.59, 1.01) |
| 0–4 | 0.27 (0.01, 1.53) | 0.54 (0.07, 1.97) | 0.27 (0.01, 1.52) | 0.54 (0.07, 1.96) | 0.54 (0.07, 1.95) | 0.00 (0.00, 0.00) |
| 5–11 | 0.37 (0.05, 1.35) | 0.18 (0.00, 1.03) | 0.18 (0.00, 1.02) | 0.18 (0.00, 1.02) | 0.91 (0.29, 2.12) | 0.36 (0.04, 1.31) |
| 12–15 | 0.32 (0.01, 1.77) | 0.32 (0.01, 1.77) | 0.00 (0.00, 0.00) | 0.00 (0.00, 0.00) | 0.00 (0.00, 0.00) | 0.31 (0.01, 1.72) |
| 16–19 | 0.00 (0.00, 0.00) | 0.28 (0.01, 1.58) | 0.56 (0.07, 2.02) | 0.83 (0.17, 2.42) | 0.00 (0.00, 0.00) | 0.00 (0.00, 0.00) |
| 20–24 | 0.20 (0.01, 1.13) | 0.60 (0.12, 1.76) | 0.20 (0.00, 1.09) | 0.00 (0.00, 0.00) | 0.37 (0.04, 1.34) | 0.55 (0.11, 1.61) |
| 25–29 | 0.00 (0.00, 0.00) | 0.41 (0.05, 1.49) | 0.40 (0.05, 1.45) | 0.38 (0.05, 1.39) | 0.00 (0.00, 0.00) | 0.36 (0.04, 1.30) |
| 30–39 | 0.46 (0.12, 1.17) | 1.00 (0.46, 1.91) | 0.33 (0.07, 0.96) | 0.53 (0.17, 1.23) | 1.12 (0.56, 2.00) | 1.37 (0.75, 2.30) |
| 40–49 | 0.32 (0.07, 0.95) | 0.44 (0.12, 1.12) | 0.55 (0.18, 1.28) | 0.66 (0.24, 1.44) | 0.55 (0.18, 1.29) | 0.33 (0.07, 0.97) |
| 50–59 | 0.68 (0.27, 1.40) | 0.87 (0.40, 1.65) | 0.97 (0.46, 1.78) | 0.68 (0.27, 1.40) | 0.78 (0.34, 1.54) | 0.99 (0.47, 1.82) |
| 60–69 | 0.67 (0.22, 1.57) | 0.26 (0.03, 0.94) | 0.51 (0.14, 1.30) | 0.99 (0.43, 1.96) | 1.09 (0.50, 2.06) | 1.29 (0.64, 2.31) |
| 70–79 | 0.24 (0.01, 1.34) | 1.62 (0.65, 3.34) | 1.09 (0.35, 2.54) | 0.82 (0.22, 2.11) | 2.56 (1.36, 4.38) | 0.95 (0.31, 2.21) |
| ≥80 | 2.67 (0.98, 5.82) | 0.43 (0.01, 2.39) | 1.25 (0.26, 3.64) | 0.40 (0.01, 2.24) | 1.17 (0.24, 3.41) | 2.27 (0.83, 4.95) |
| **Transverse myelitis, broad definition** | | | | | | |
| **Both sexes** | | | | | | |
| All ages | 1.27 (1.09, 1.47) | 1.84 (1.63, 2.09) | 1.69 (1.48, 1.92) | 1.81 (1.60, 2.04) | 1.95 (1.73, 2.19) | 1.93 (1.71, 2.17) |
| 0–4 | 0.56 (0.15, 1.44) | 0.56 (0.15, 1.43) | 0.28 (0.03, 1.01) | 0.83 (0.31, 1.82) | 0.69 (0.23, 1.62) | 0.28 (0.03, 1.00) |
| 5–11 | 0.57 (0.21, 1.24) | 0.56 (0.21, 1.23) | 0.56 (0.21, 1.22) | 0.46 (0.15, 1.08) | 0.74 (0.32, 1.46) | 0.56 (0.20, 1.21) |
| 12–15 | 0.82 (0.26, 1.90) | 0.97 (0.36, 2.12) | 0.81 (0.26, 1.89) | 0.64 (0.18, 1.65) | 0.80 (0.26, 1.86) | 1.42 (0.65, 2.69) |
| 16–19 | 0.88 (0.32, 1.91) | 1.76 (0.91, 3.07) | 1.01 (0.41, 2.09) | 1.00 (0.40, 2.06) | 2.57 (1.52, 4.06) | 1.01 (0.41, 2.08) |
| 20–24 | 0.53 (0.17, 1.23) | 1.57 (0.88, 2.59) | 1.33 (0.71, 2.28) | 1.29 (0.69, 2.21) | 1.94 (1.18, 2.99) | 1.35 (0.74, 2.26) |
| 25–29 | 1.62 (0.91, 2.67) | 2.11 (1.29, 3.26) | 2.26 (1.42, 3.43) | 2.08 (1.29, 3.18) | 1.81 (1.09, 2.83) | 1.76 (1.06, 2.75) |
| 30–39 | 1.50 (0.99, 2.19) | 2.57 (1.89, 3.42) | 1.99 (1.40, 2.74) | 2.30 (1.67, 3.09) | 2.28 (1.66, 3.05) | 2.75 (2.08, 3.57) |
| 40–49 | 1.86 (1.29, 2.58) | 1.82 (1.26, 2.54) | 2.31 (1.67, 3.12) | 2.64 (1.96, 3.49) | 1.94 (1.36, 2.69) | 2.64 (1.96, 3.50) |
| 50–59 | 1.45 (0.98, 2.07) | 2.40 (1.78, 3.16) | 2.21 (1.62, 2.95) | 2.46 (1.83, 3.24) | 2.04 (1.47, 2.76) | 2.35 (1.73, 3.12) |
| 60–69 | 0.90 (0.49, 1.52) | 1.69 (1.11, 2.46) | 1.84 (1.24, 2.62) | 1.73 (1.16, 2.49) | 2.15 (1.52, 2.97) | 2.04 (1.43, 2.82) |
| 70–79 | 1.68 (0.94, 2.77) | 2.48 (1.57, 3.72) | 1.82 (1.08, 2.88) | 1.92 (1.17, 2.97) | 2.94 (2.01, 4.15) | 2.03 (1.29, 3.04) |
| ≥80 | 2.08 (1.07, 3.63) | 2.02 (1.04, 3.53) | 1.48 (0.68, 2.81) | 1.60 (0.77, 2.94) | 2.65 (1.54, 4.24) | 2.29 (1.28, 3.77) |
| **Females** | | | | | | |
| All ages | 1.51 (1.23, 1.83) | 2.39 (2.04, 2.78) | 2.06 (1.74, 2.42) | 2.14 (1.82, 2.50) | 2.12 (1.80, 2.48) | 2.23 (1.90, 2.59) |
| 0–4 | 0.57 (0.07, 2.07) | 0.28 (0.01, 1.59) | 0.29 (0.01, 1.59) | 0.86 (0.18, 2.50) | 0.57 (0.07, 2.06) | 0.57 (0.07, 2.05) |
| 5–11 | 0.39 (0.05, 1.40) | 0.57 (0.12, 1.68) | 0.38 (0.05, 1.38) | 0.57 (0.12, 1.66) | 0.57 (0.12, 1.65) | 0.38 (0.05, 1.37) |
| 12–15 | 0.67 (0.08, 2.42) | 1.33 (0.36, 3.40) | 0.99 (0.20, 2.90) | 0.99 (0.20, 2.88) | 0.65 (0.08, 2.34) | 1.28 (0.35, 3.28) |
| 16–19 | 1.21 (0.33, 3.10) | 2.42 (1.04, 4.77) | 1.49 (0.49, 3.49) | 0.88 (0.18, 2.58) | 3.81 (2.03, 6.52) | 1.77 (0.65, 3.86) |
| 20–24 | 0.88 (0.24, 2.26) | 1.75 (0.76, 3.45) | 1.71 (0.74, 3.37) | 2.08 (1.00, 3.83) | 2.03 (0.97, 3.74) | 1.81 (0.83, 3.44) |
| 25–29 | 2.19 (1.05, 4.03) | 3.24 (1.81, 5.35) | 3.59 (2.09, 5.74) | 2.85 (1.56, 4.78) | 2.56 (1.36, 4.38) | 2.30 (1.19, 4.02) |

| | | | | | | |
|---|---|---|---|---|---|---|
| 30–39 | 1.85 (1.08, 2.96) | 3.55 (2.44, 4.98) | 2.44 (1.55, 3.66) | 2.81 (1.85, 4.08) | 2.53 (1.64, 3.74) | 3.06 (2.08, 4.34) |
| 40–49 | 2.40 (1.52, 3.60) | 2.73 (1.78, 4.00) | 3.16 (2.13, 4.51) | 3.48 (2.39, 4.88) | 2.52 (1.62, 3.76) | 3.78 (2.65, 5.24) |
| 50–59 | 1.34 (0.73, 2.25) | 2.86 (1.93, 4.09) | 2.67 (1.78, 3.87) | 2.68 (1.78, 3.88) | 2.21 (1.40, 3.32) | 2.62 (1.73, 3.81) |
| 60–69 | 1.12 (0.51, 2.13) | 2.29 (1.38, 3.58) | 2.13 (1.26, 3.36) | 1.73 (0.97, 2.86) | 2.03 (1.20, 3.20) | 1.76 (1.00, 2.85) |
| 70–79 | 2.71 (1.44, 4.64) | 2.42 (1.25, 4.23) | 1.71 (0.78, 3.24) | 1.62 (0.74, 3.08) | 2.75 (1.57, 4.47) | 2.15 (1.14, 3.67) |
| ≥80 | 1.42 (0.46, 3.30) | 2.49 (1.14, 4.73) | 0.81 (0.17, 2.38) | 1.86 (0.75, 3.84) | 1.82 (0.73, 3.75) | 2.04 (0.88, 4.02) |
| **Males** | | | | | | |
| All ages | 1.02 (0.80, 1.29) | 1.29 (1.03, 1.59) | 1.31 (1.06, 1.61) | 1.47 (1.20, 1.78) | 1.78 (1.49, 2.12) | 1.62 (1.34, 1.94) |
| 0–4 | 0.55 (0.07, 1.98) | 0.82 (0.17, 2.39) | 0.27 (0.01, 1.52) | 0.81 (0.17, 2.38) | 0.81 (0.17, 2.37) | 0.00 (0.00, 0.00) |
| 5–11 | 0.75 (0.20, 1.91) | 0.55 (0.11, 1.62) | 0.73 (0.20, 1.88) | 0.36 (0.04, 1.32) | 0.91 (0.29, 2.12) | 0.73 (0.20, 1.86) |
| 12–15 | 0.95 (0.20, 2.79) | 0.64 (0.08, 2.30) | 0.63 (0.08, 2.29) | 0.32 (0.01, 1.76) | 0.94 (0.19, 2.74) | 1.54 (0.50, 3.60) |
| 16–19 | 0.57 (0.07, 2.05) | 1.14 (0.31, 2.91) | 0.56 (0.07, 2.02) | 1.11 (0.30, 2.83) | 1.39 (0.45, 3.24) | 0.28 (0.01, 1.57) |
| 20–24 | 0.20 (0.01, 1.13) | 1.41 (0.57, 2.90) | 0.98 (0.32, 2.29) | 0.57 (0.12, 1.66) | 1.85 (0.89, 3.40) | 0.92 (0.30, 2.15) |
| 25–29 | 1.06 (0.35, 2.48) | 1.03 (0.34, 2.41) | 1.00 (0.33, 2.34) | 1.35 (0.54, 2.78) | 1.11 (0.41, 2.42) | 1.26 (0.51, 2.59) |
| 30–39 | 1.14 (0.55, 2.10) | 1.56 (0.85, 2.62) | 1.53 (0.83, 2.56) | 1.80 (1.05, 2.87) | 2.03 (1.24, 3.14) | 2.45 (1.59, 3.62) |
| 40–49 | 1.29 (0.67, 2.26) | 0.87 (0.38, 1.72) | 1.43 (0.76, 2.44) | 1.77 (1.01, 2.87) | 1.33 (0.69, 2.32) | 1.44 (0.77, 2.47) |
| 50–59 | 1.55 (0.89, 2.53) | 1.93 (1.18, 2.98) | 1.74 (1.03, 2.75) | 2.24 (1.42, 3.36) | 1.86 (1.12, 2.90) | 2.08 (1.29, 3.17) |
| 60–69 | 0.67 (0.22, 1.57) | 1.04 (0.45, 2.05) | 1.53 (0.79, 2.67) | 1.74 (0.95, 2.91) | 2.29 (1.38, 3.58) | 2.34 (1.43, 3.62) |
| 70–79 | 0.48 (0.06, 1.74) | 2.55 (1.27, 4.56) | 1.96 (0.89, 3.71) | 2.27 (1.13, 4.06) | 3.15 (1.80, 5.12) | 1.89 (0.91, 3.48) |
| ≥80 | 3.12 (1.25, 6.43) | 1.29 (0.27, 3.77) | 2.49 (0.92, 5.43) | 1.21 (0.25, 3.53) | 3.89 (1.87, 7.16) | 2.65 (1.07, 5.47) |
